# Sample size estimates for biomarker-based outcome measures in clinical trials in autosomal dominant Alzheimer’s disease

**DOI:** 10.1101/2024.11.12.24316919

**Authors:** David M Cash, Katy E Morgan, Antoinette O’Connor, Thomas D Veale, Ian B Malone, Teresa Poole, Tammie LS Benzinger, Brian A Gordon, Laura Ibanez, Yan Li, Jorge J. Llibre-Guerra, Eric McDade, Guoqiao Wang, Jasmeer P Chhatwal, Gregory S Day, Edward Huey, Mathias Jucker, Johannes Levin, Yoshiki Niimi, James M Noble, Jee Hoon Roh, Racquel Sánchez-Valle, Peter R Schofield, Randall J Bateman, Chris Frost, Nick C Fox, The Dominantly Inherited Alzheimer Network (DIAN)

## Abstract

**INTRODUCTION:** Alzheimer disease (AD)-modifying therapies are approved for treatment of early-symptomatic AD. Autosomal dominant AD (ADAD) provides a unique opportunity to test therapies in presymptomatic individuals.

**METHODS:** Using data from the Dominantly Inherited Alzheimer Network (DIAN), sample sizes for clinical trials were estimated for various cognitive, imaging, and CSF outcomes. Sample sizes were computed for detecting a reduction of either absolute levels of AD-related pathology (amyloid, tau) or change over time in neurodegeneration (atrophy, hypometabolism, cognitive change).

**RESULTS:** Biomarkers measuring amyloid and tau pathology had required sample sizes below 200 participants per arm (examples CSF Aβ42/40: 47[95%CI 25,104], cortical PIB 49[28,99], CSF p-tau181 74[48,125]) for a four-year trial in presymptomatic individuals (CDR=0) to have 80% power (5% statistical significance) to detect a 25% reduction in absolute levels of pathology, allowing 40% dropout. For cognitive, MRI, and FDG, it was more appropriate to detect a 50% reduction in rate of change. Sample sizes ranged from 250-900 (examples hippocampal volume: 338[131,2096], cognitive composite: 326[157,1074]). MRI, FDG and cognitive outcomes had lower sample sizes when including indivduals with mild impairment (CDR=0.5 and 1) as well as presymptomatic individuals (CDR=0).

**DISCUSSION:** Despite the rarity of ADAD, presymptomatic clinical trials with feasible sample sizes given the number of cases appear possible.

## 1. Introduction

After many unsuccessful trials involving potential disesase modifying therapies (DMT) for Alzheimer’s disease (AD), recent trials of anti-amyloid treatments[1–3] provide much-needed hope to patients and their families. These DMTs substantially reduced amyloid plaques and slowed cognitive decline compared to placebo in early symptomatic AD[1–3]. As amyloid pathology begins decades before symptom onset[4,5], anti-amyloid DMTs may show the greatest benefit when administered earlier in the disease course before downstream pathological processes gain momentum leading to the onset of symptoms and irreversible neurodegeneration. There are other classes of therapies being developed as well, which may also benefit from early intervention in individuals who are unimpaired or have early impairment.

An effective therapy is urgently needed for individuals with Autosomal Dominant forms of AD (ADAD), a rare form comprising less than 1% of all cases. ADAD is caused by the presence of pathogenic mutations in the Presenilin 1 (*PSEN1*), Presenilin 2 (*PSEN2*) or Amyloid Precursor Protein (*APP*) genes[6]. These mutations are nearly 100% penetrant with a reasonably consistent age at onset within families[7] that typically occurs decades earlier than sporadic AD. Thus, ADAD provides a unique opportunity to test DMTs in the early stages of disease, including presymptomatic carriers, who will almost certainly develop symptoms within a predictable time window and who are highly motivated to participate in trials. Identifying successful treatments in presymptomatic ADAD could increase confidence of efficacy in the biomarker-positive, cognitively unimpaired phase of sporadic AD. However, how to best assess treatment efficacy during this window is not straightforward. Clinical “prevention” trials present design challenges that include identifying appropriate participants, determining meaningful endpoints that are sensitive to change, and powering the trial adequately in terms of enrolment and duration[8,9]. Biomarkers can measure different aspects of AD and could be useful as potential outcomes in trials involving individuals prior to symptom onset. Some biomarkers reflect increased levels of amyloid plaques and neurofibrillary tau tangles, the primary pathologies that define AD and are the target for removal by many DMTs. Recent FDA guidelines[10] have endorsed amyloid biomarkers as outcome measures in trials involving participants where AD pathology is present but cognitive impairment is either absent or subtle. If these therapies are effective, then biomarker levels related to amyloid and tau burden should return towards normal levels. Other measures, such as brain volume and cognitive function, reflect downstream changes caused by neurodegeneration and are usually considered irreversible. For these kinds of biomarkers, progession measured through rate of change (e.g. brain atrophy in % loss/year) is more clinically relevant rather than the absolute level.

Biomarker-based evidence of treatment-related reductions in pathological burden, and a clinically meaningful outcome are both required to demonstrate a disease-modifying effect in a classical parallel arm designed randomised controlled trial. When performing prevention trials in a rare population like ADAD, the right design is essential to minimize the number of individuals needed to detect a clinically significant treatment effect with the desired statistical power over a feasible trial duration. There is no clear consensus for a trial duration in presymptomatic trials. A duration that is too short would require too many participants to detect a biological or clinical effect. Durations that are too long would raise concerns about ethics, safety, cost, and participant withdrawal. Recent presymptomatic trials [11,12] have proposed a trial duration of four years.

In this study, we used observational study data from the Dominantly Inherited Alzheimer’s Network observational study (DIAN-OBS), a large multicentre study of ADAD, to estimate sample sizes for prospective prevention trials in ADAD. Target treatment effects for these estimates were defined based on the type of outcome measure. For candidate outcomes reflecting primary pathologies, such as amyloid positron emission tomography (PET) or soluble measures of cerebrospinal fluid (CSF) amyloid, phospho-tau 181 (p-tau181) and total tau, we estimated sample sizes required to detect a reduction in the absolute level by the end of a four-year trial. For outcomes that reflect downstream neurodegeneration (cognitive scales, FDG PET and volumetric magnetic resonance imaging (MRI)), sample sizes were based on detecting a reduction in rates of change over time.

## 2. Methods

### 2.1 Participants

All data came from participants enrolled in DIAN-OBS - a worldwide, multi-modal study of ADAD mutation carriers and non-carrier family members[13], who serve as a valuable environmentally similar control group, enabling characterisation of the divergence of disease-related changes from normal aging.

DIAN-OBS was designed to parallel clinical trials: integrating rigorously collected, longitudinal data across multiple centres and including a wide array of imaging, fluid biomarker, and clinical measures. Indeed, the clinical trial DIAN-TU-001 (ClinicalTrials.gov Identifier: NCT04623242, NCT01760005) included many individuals from DIAN-OBS, allowing for the potential of a run-in phase as part of their design[12,14]. Detailed information concerning the DIAN-OBS study protocol, including MRI and PET image acquisition, has been reported previously[15].

Participants in DIAN-OBS are from families known to carry a pathological mutation in *PSEN1*, *PSEN2*, or *APP* genes. Data were taken from the 14^th^ semi-annual data freeze (2020), which included cognitive, biomarker and imaging data from 534 participants, 372 of whom have longitudinal data. Since age at symptomatic onset is relatively consistent within ADAD families [7] and most mutation carriers in DIAN-OBS are presymptomatic at enrolment, estimated years to expected symptom onset (EYO) can be calculated for individual participants[7] by subtracting the age their affected parent first developed symptoms from the participant’s age at their visit. In this way, EYO can be used to determine eligibility for trials amongst presymptomatic mutation carriers. While EYO is not clinically relevant for the non-carriers, estimating it helps ensure that the non-carrier group is demographically similar to the carrier group and helps to account for any age-related changes.

From this data freeze, we identified participants who would be elgible for a putative presymptomatic trial according to the following criteria: (1) an EYO from -15 to +10 years (i.e., between 15 years before and up to 10 years after predicted onset), which is the same range as used in the DIAN-TU-001 trial, and (2) a global Clinical Dementia Rating® (CDR) scale[16] of 0 (i.e. cognitively unimpaired). 178 (90 carriers and 88 non-carriers) of the 372 participants with longitudinal data had a visit that would satisfy these criteria as well as having at least one subsequent follow-up. We refer to these 90 carriers as “presymptomatic trial-eligible” participants. In addition, we looked at trials where the CDR eligibility criterion was relaxed to include those with mild impairment (CDR=0 to 1 inclusive), as this provides a benchmark to the recently completed DIAN-TU-001 study. 244 of the participants (the 88 non-carriers together with 156 carriers) met these expanded criteria.

### 2.2 Informed Consent

The DIAN-OBS study was reviewed and approved by the appropriate Institutional Review Boards and research ethics committees for each participating site. Informed consent was obstained from all participants.

### 2.3 Image Processing

MRI images were processed by the central imaging core at Washington University using FreeSurfer 5.3[17] as well as an in-house whole brain parcellation technique based on Geodesic Information Flow[18]. For bilateral structures, left and right volumetric measurements were summed. Total intracranial volume (TIV) was also extracted from the T1 image using Statistical Parametric Mapping 12 (SPM12) and served as a proxy for head size[19]. Direct measures of whole brain and ventricular atrophy were calculated using the boundary shift integral (BSI)[20–22]. Follow-up data acquired on different MRI scanners (58 scans from 40 participants) from their first visit were excluded as were data (40 scans from 26 participants) with significant motion, geometric distortion between timepoints, and non-AD pathology (infarcts, traumatic injury).

^18^F-flouroxyglucose (FDG) and ^11^C-Pittsburgh compound B (PIB) PET images, measuring glucose metabolism and amyloid accumulation respectively, were processed using the PET Unified Pipeline (PUP) pipeline[23] that provides regional Standard Uptake Value Ratio (SUVR) measures for all FreeSurfer cortical regions of interest (ROIs), where the whole cerebellum served as the reference region. As there has been evidence in some cases of amyloid deposition in the cerebellum in ADAD[24,25], we examined SUVRs with the brainstem as a reference, but there was minimal change in the results. SUVR values were obtained with partial volume correction (PVC) using the geometric transfer matrix approach[26,27], as well as without PVC. The PVC PIB SUVR values were used for sample size analysis as they consistently produced greater differences in mean levels between carriers and non-carriers than the non-PVC PIB values. In contrast, the non-PVC FDG SUVR produced greater differences in mean levels between carriers and non-carriers than the PVC, so these values were included in the sample size analysis instead for the FDG outcome measure. As with the MRI biomarkers, scans acquired on different PET scanners (52 PIB scans from 35 participants and 48 FDG scans from 34 participants) were excluded from analysis.

### 2.4 Cerebrospinal fluid analysis

Collection of CSF was performed according to a protocol consistent with Alzheimer’s Disease Neuroimaging Initiative (ADNI) and analysis was performed by the central biomarker core at Washington University[28]. For this study, we included the Aβ 1-40, Aβ 1-42, p-tau181 and total tau measures from two immunoassays: the Luminex bead-based multi-plexed xMAP technology (INNO-BIA AlzBio3, Innogenetics) and the Lumipulse automated immunoassay system (LUMIPULSE G1200; Fujirebio, Malvern, PA, USA). For the CSF XMAP, both the cross-sectional and longitudinal processing (CSF XMAP LONG) pipelines were considered.

### 2.5 Choosing a target therapeutic effect

An important decision for sample size estimation is how to define a target treatment effect. The specified treatment effect should be large enough to represent a clinically meaningful benefit, but not so large that it would be implausible to achieve and result in the trial being underpowered.

Treatment effects are often expressed relative to what a “completely successful” treatment would achieve. In defining such a treatment effect, we believe that there needs to be a distinction between measures of primary pathology (PIB PET and CSF) and outcomes measuring downstream processes reflecting neurodegeneration (MRI, FDG PET, cognitive). For biomarkers of downstream processes, a treatment would be judged completely successful if it were to reduce the average *rate of change* in the biomarker to that observed in normal ageing. This is because current treatments are not (yet) expected either to reverse the course of neurodegeneration (e.g. to restore lost neurones) or to be able also to halt losses associated with normal ageing processes in individuals with no biomarker evidence of AD pathology. However, for biomarkers of primary pathology, there is growing evidence that slowing the rate of pathological accumulation in amyloid and tau will not be sufficient to modify the disease in manner that would be clinically meaningful to patients. Rather, large reductions in the absolute levels of primary pathology by the end of the study would be needed in order to provide a tangible, clinical benefit to patients[29]. In fact, results from recent trials of anti-amyloid therapies have shown it is possible to achieve very substantial reductions in PET and CSF amyloid outcomes[1,3,12,30], and in some cases also to show slowing of cognitive decline[1,2]. Therefore, for PIB PET and CSF outcome measures an effect on amyloid burden would be considered completely (100%) successful if average *absolute levels* were reduced to normal by the end of the study.

Other treatment effects can be defined relative to a completely successful one. For example, a 50% effective treatment acting on a marker of neurodegeneration would halve the average excess rate of change (over and above that seen in normal ageing) whereas a 50% effective treatment acting on a measure of amyloid burden would halve the average excess level. Deciding on a clinically relevant target treatment effect is not straightforward. For markers of neurodegeneration we chose a 50% reduction in the rate of change in carriers, relative to the rate of change in non-carriers (Figure 1, left panel). A reduction of this magnitude is similar to one thought to be clinically meaningful based on decline in the Preclinical Alzheimer’s Cognitive Composite (PACC) in Aβ+ cognitively normal patients compared to those that were Aβ-[9]. For measures of amyloid burden, we chose a reduction of 25% in excess level (level over and above that in non-carriers; Figure 1, right panel) by the end of the trial, as this would likely be the minimum level that could provide clinical benefit[31] for presymptomatic individuals, who are likely to have lower levels of amyloid burden in comparison to individuals enrolled in current phase III trials. While phase III clinical trials of anti-amyloid DMTs have have shown far greater reductions in amyloid PET, our proposed level of 25% reduction has been observed in amyloid PET for some ADAD studies[32] and may be more in line with what is observed in CSF and plasma biomarkers. Basing sample size calculations on reductions of greater than 25% might mean that clinically important reductions are missed. Further, if reductions are in truth larger than 25% then basing the sample size on a 25% reduction will be conservative, and the trial will have increased statistical power to demonstrate statistically significant effects. Further details on the definition of the target therapeutic effects are given in the Statistical Appendix found in the Supplementary Material.

**Figure 1.**
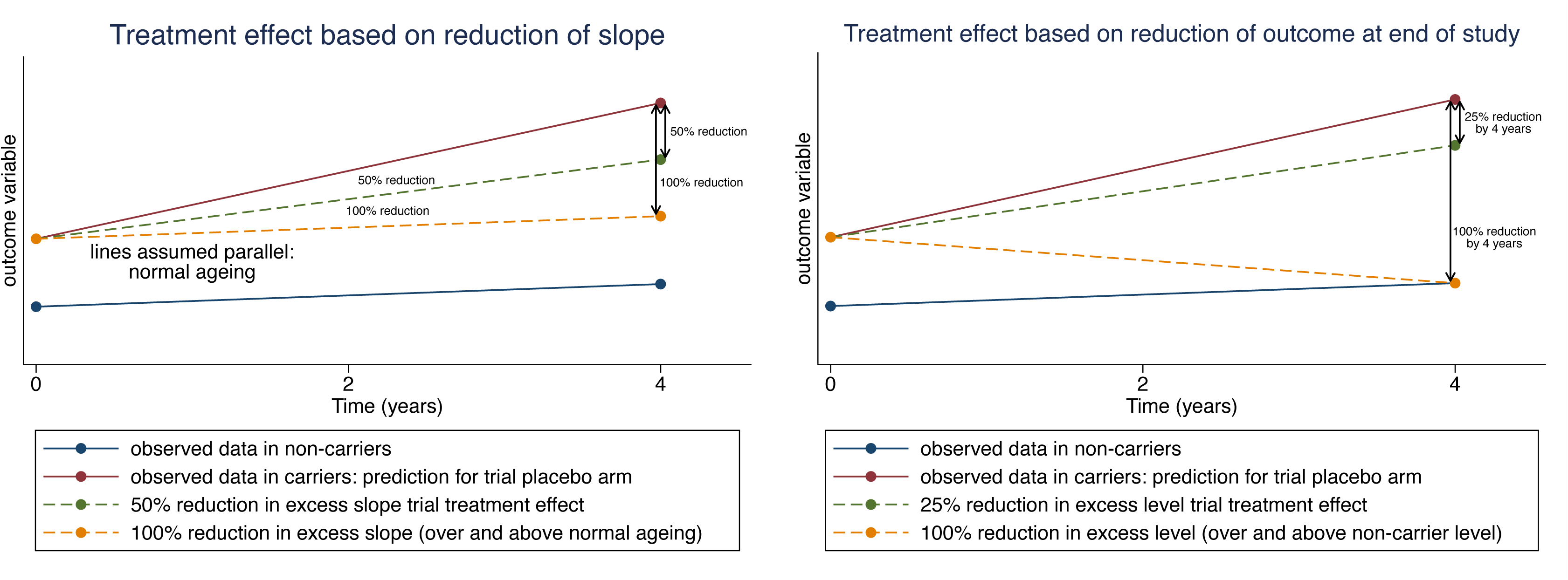
Visual examples of the two types of treatment effects considered in this sample size analysis. (Left) The treatment effect is based on a reduction of the slope, with a 100% therapeutic effect being defined as one that reduces the slope in the treatment arm to the slope observed in the non-carriers. The outcome variable may undergo a transformation before analysis so that trajectories are more linear in nature. (Right) The treatment effect is based on reducing the outcome measure by the end of the study, with a 100% therapeutic effect defined as one that would reduce the absolute level of the outcome measure to the expected outcome in non-carriers. While the analysis may be conducted on the log transformed scale, the treatment effect is defined on the scale of the original outcome variable in order to make the definition of the treatment effect more interpretable.

### 2.6 Study Design and Statistical Methods

The methodology for assessing the trial designs is described in full in the Statistical Appendix in the Supplementary Material. Briefly, a two-stage approach was used[33,34]. In stage 1, linear mixed models (LMM) were fitted to the observational repeated measures data from carriers and non-carriers in DIAN-OBS to obtain estimates of parameters that allow us to define plausible target therapeutic effects and to quantify components of variability. In stage 2, estimates from the LMM are used to compute sample size requirements for four-year trials with single measures of candidate outcome measures at baseline and follow-up (or a single direct measure of four-year change).

We selected candidate outcome measures from the ADAD literature; ideal outcome measures are sensitive biomarkers that reflect the key disease processes[17,28,35–37]. We selected PIB SUVR from six ROIs (precuneus, posterior cingulate, inferior parietal, interior temporal, middle temporal, and a mean cortical SUVR of the precuneus, prefrontal cortex, gyrus rectus, and lateral temporal regions) and six CSF measures (Aβ1–40, Aβ1–42, total tau, p-tau181, the Aβ1–42 to 1–40 ratio, and the p-tau181 to Aβ1-42 ratio) as measures of AD-related pathology. CSF total tau is included in this group although it is regarded variably as a marker of AD pathology and neurodegeneration. For the MRI measures of brain atrophy, volumes from five ROIs (whole brain, lateral ventricle, hippocampus, precuneus, and posterior cingulate), and two direct measures of change (brain and lateral ventricle BSI) were used. Measures of hypometabolism were assessed using six ROIs from FDG PET (precuneus, posterior cingulate, inferior parietal, hippocampus, banks of superior temporal sulcus (banks STS), and a mean cortical SUVR). Finally, cognitive decline was measured from the mini mental state exam (MMSE), the CDR sum-of-box scores (CDR-SB), and a cognitive composite including four scales: MMSE, the Logical Memory delayed recall score from the Weschler Memory Scale-Revised, animal naming, and the digit symbol score substitution from the Wechsler Adult Intelligence scale-Revised. To generate this composite, each scale was separately z-scored, and the mean of the four z-scores was taken. This composite is designed to capture earlier changes and is very similar to the PACC, as well as composites used in DIAN-OBS[38,39] and DIAN-TU[40]. MRI, PET, and CSF markers were log-transformed to provide outcome measures on a scale representing annual percentage change from baseline. Cognitive scores were left untransformed, as this is common practice in phase 3 trials, and it allows a more intuitive interpretation of the resulting treatment effects.

We considered putative placebo-controlled two-arm parallel trials (1:1 randomisation) with a duration of four years. While a trial duration of four years tends to be longer than most phase 3 trials in sporadic AD, a longer duration is likely needed for studies involving presymptomatic participants to allow changes in the outcome that can be detected. This duration also matches the “common close” design of DIAN-TU-001, where all participants were followed up for at least four years. For all outcomes other than “direct” measures of change we assumed that the outcome measure would be obtained at baseline (pre-randomization) and then again at the end of the follow-up period (four years). Based on this design, we assumed that an analysis of covariance (ANCOVA) would be used for the statistical analysis, with MRI, PET and CSF markers log-transformed as in the analysis of DIAN-OBS. For volumetric MRI measures we assumed that TIV would be included as an additional covariate in the ANCOVA model, as TIV serves as a proxy for head size. For “direct” measures of change between the two timepoints, such as those obtained from the BSI, we assumed that between group comparisons would be carried out using a t-test. In addition to considering trial designs where presymptomatic (global CDR score of 0, i.e. cognitively normal) participants from DIAN-OBS would be eligible, we also considered a trial design similar to DIAN-TU-001, enrolling all participants with a global CDR score of 0 to 1, inclusive.

To define target theraupeutic effects for these trial designs, estimates of the mean rates of change over time in these candidate outcome measures for both carriers and non-carriers are needed, as well as estimates of relevant variances and covariances in carriers. To obtain estimates of these key parameters, we fit LMMs (see Supplementary Materials for details) to the longitudinal data from the full sample and presymptomatic subset of DIAN-OBS. Data from DIAN-OBS participants were included if there were outcome measures available at both a “baseline” visit that satisfied the specified eligibility criteria and at least one subsequent follow-up visit. For volumetric MRI measures, TIV and its interaction with time were included in the LMM. We excluded outcome measures from sample size calculation when the LMM did not converge, or when they were not considered to be suitable candidates for a future trial (see Supplementary Materials for more details).

We assumed 40% dropout rate at 4 years in our putative trials. This is slightly conservative compared to recent clinical trials: 27% over four years for DIAN-TU-001 trials of gantenerumab and solanezumab[12], 23% for the TRAILBLAZER-ALZ-2 randomized controlled trial (RCT) of donanemab[2], 17% of 18 months for the Clarity AD RCT of lecanemab[1], and 29% for 4.5 years in the A4 study of solanezumab[41]. For all outcome measures, we obtained sample size estimates that would be required to detect a clinically meaningful benefit with 80% statistical power using a two-sided significance level of 5%. Uncertainty in the resulting sample size estimates was quantified using 95% bias-corrected and accelerated (BCa) confidence intervals obtained through bootstrapping. A modified version of the Stata package slopepower[42] was used to implement the sample size calculations (modifications including allowing adjustment for TIV and analysis of direct measures of change).

## 3. Results

Table 1 shows baseline demographics for trial-eligible participants included in the analysis. 90 carriers would be eligible for a presymptomatic trial, while 156 would be eligible for trials that also enrolled individuals with mild impairment (CDR=0-1). Non-carriers and carriers were well-matched for sex, age, and EYO. There were four non-carriers with a baseline global CDR of 0.5 (very mild impairment); three reverted to a global CDR of 0 at subsequent visits. Most individuals came from *PSEN1* families. The amount of data available for sample size analysis depended on the modality, with the number of participants and number of observations included in the analysis by modality also listed in Table 1. Cognitive variables had the most data, followed by CSF and MRI, and finally PET biomarkers.

**Table 1.** Baseline demographics of all participants included in the analysis. A participant was considered trial eligible if they met the criteria at one “baseline” visit and at least one subsequent visit. There were five carriers who had a global CDR score greater than 0 at an earlier visit but reverted to a CDR global score of 0 at a subsequent visit and had longitudinal data so that they could be included in the presymptomatic-only trial eligible population.

|  | Non-carriers | Carriers<br>(CDR=0) | Carriers<br>(CDR= 0 – 1) |
| --- | --- | --- | --- |
| Number of participants | 88 | 90 | 156 |
| Sex, N female (%) | 48 (55%) | 60 (67%) | 92 (59%) |
| Age (years), mean<br>(SD) | 41.2 (7.6) | 38.8 (8.1) | 41.6 (9.0) |
| EYO (years), mean<br>(SD) | -5.7 (6.6) | -6.9 (5.3) | -3.7 (6.4) |
| CDR global (0/0.5/1) | 84/4/0 | 90/0/0 | 85/54/17 |
| Family mutation, N<br>(*PS1/PS2/APP*) | 55/8/25 | 67/5/18 | 121/6/29 |
| Participants<br>(Observations) with: |  |  |  |
| Cognitive data | 88 (248) | 90 (259) | 156 (470) |
| Structural MRI | 65 (173) | 60 (164) | 106 (292) |
| PIB PET | 46 (124) | 50 (124) | 80 (201) |
| FDG PET | 50 (131) | 48 (123) | 90 (227) |
| CSF XMAP | 61 (157) | 61 (159) | 107 (293) |
| CSF XMAP LONG | 36 (85) | 39 (98) | 71 (185) |
| CSF LUMIPULSE | 63 (166) | 64 (174) | 113 (320) |

Based on the LMM, Figure 2 provides estimated means and 95% CIs for selected outcomes at study start and end (all outcome measures, except for BSI-based measures, are shown in Supplementary Figure 1 and Supplementary Table 1). The LMM did not converge for the following outcomes and trial scenarios: CDR-SB (CDR=0), all cross-sectional CSF XMAP measures (both) except for total tau, longitudinal CSF XMAP measures of p-tau181(CDR=0) and p-tau181/Aβ1-42(both), and FDG cortical mean SUVR (CDR=0). These were excluded from subsequent analysis. In addition, the following outcome scenarios were excluded in trials involving CDR=0 carriers as there was insufficient evidence of a substantial difference in slope between carriers and non-carriers (see Supplemenatary Material): whole brain volume, posterior cingulate volume, and all FDG measures. The FDG SUVR for posterior cingulate and hippocampus also had insufficient evidence for the CDR=0-1 trial sample. For most biomarkers, the trajectories of non-carriers over the duration of the trial remained essentially flat, reflecting that there was no evidence from the LMM that the mean rates of change were non-zero. Exceptions were a slight improvement observed on the cognitive composite (presumably practice effects), and slight decreases in cortical FDG SUVR, MRI volumes and BSI, which are expected in normal ageing. There was also a slightly negative rate of change in CSF p-tau181 and Aβ1-40 in non-carriers, as well as for carriers when using the XMAP assay. However, the LMMs did provide statistically significant evidence of differences between carriers and non-carriers, both at baseline and by the end of the proposed four-year trial duration. In most cases, these differences were greater by the end of the proposed four-year trial.

**Figure 2.**
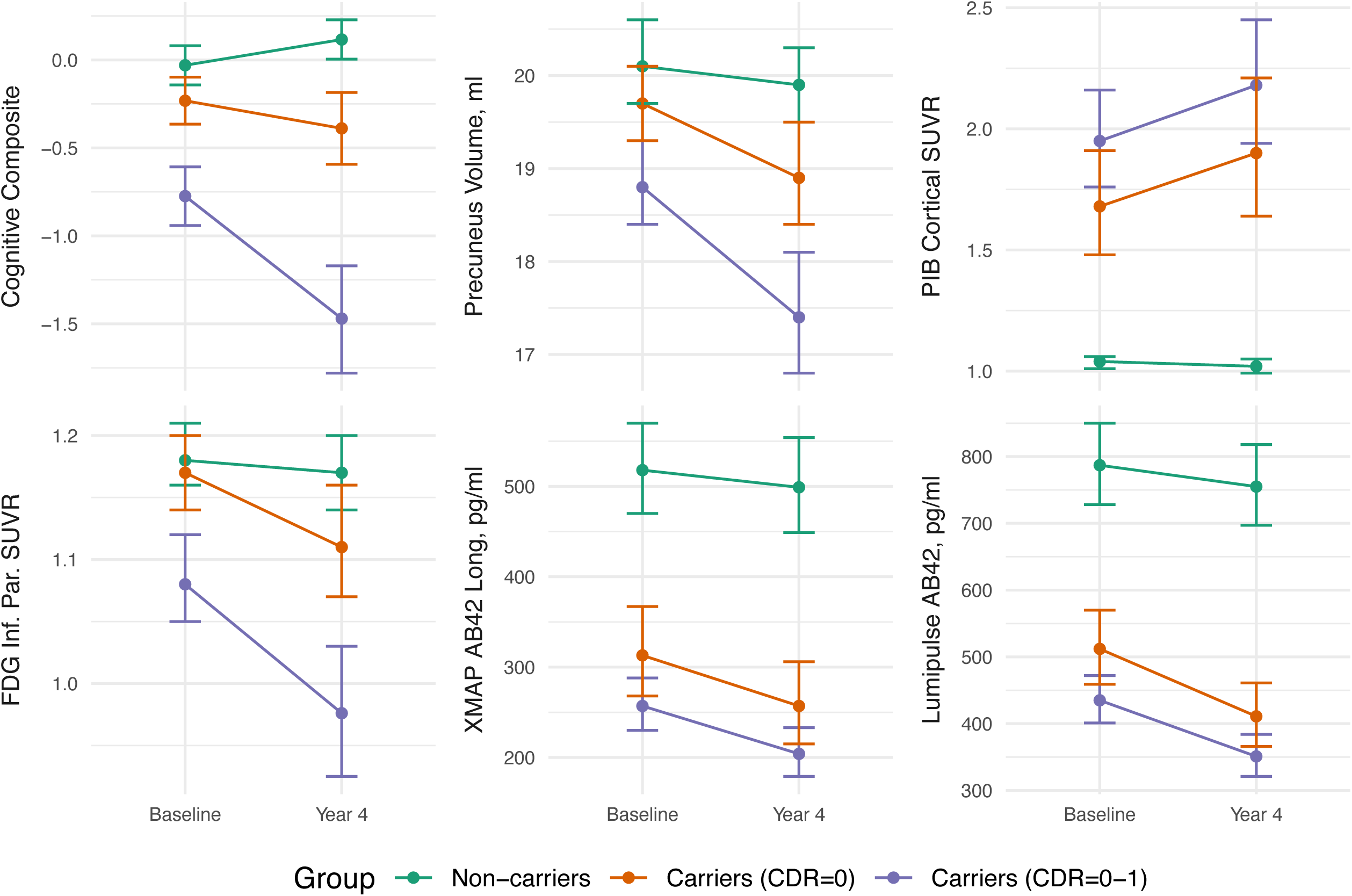
Estimated measures from selected outcomes at baseline and at four-year follow-up (means and 95% confidence intervals) for carriers and non-carriers. These estimates are based on the parameters from fitting the linear mixed effects model to participants in the observational studies who match the trial eligibility criteria. These estimates form the basis of the subsequent sample size estimates. All estimates have been back-transformed, when necessary, to plot the estimated outcome measures on the original scale. Plots for all outcomes can be observed in Supplementary Figure 1.

From the LMM estimates, the target treatment effect for 50% slowing of neurodegeneration represents a slowing of the hippocampal atrophy rate from 0.65% per year to 0.38% per year. The treatment effect of 25% reduction in mean cortical PIB PET represents a decrease from 1.90 (34 centiloids) in the placebo to 1.68 (25 centiloids) at the end of a four year trial.

The sample size estimates (with 95% BCa confidence intervals) needed to detect a treatment effect of 50% reduction in the rate of neurodegeneration (atrophy, cognitive decline, hypometabolism on FDG PET) compared to non-carriers over a four-year trial, assuming 40% dropout, with 5% significance and 80% power are shown in Figure 3 and Supplementary Table 2. For most of these candidate outcome measures, sample sizes were larger for the presymptomatic trials than for trials that would include both unimpaired and mildly impaired participants.

**Figure 3.**
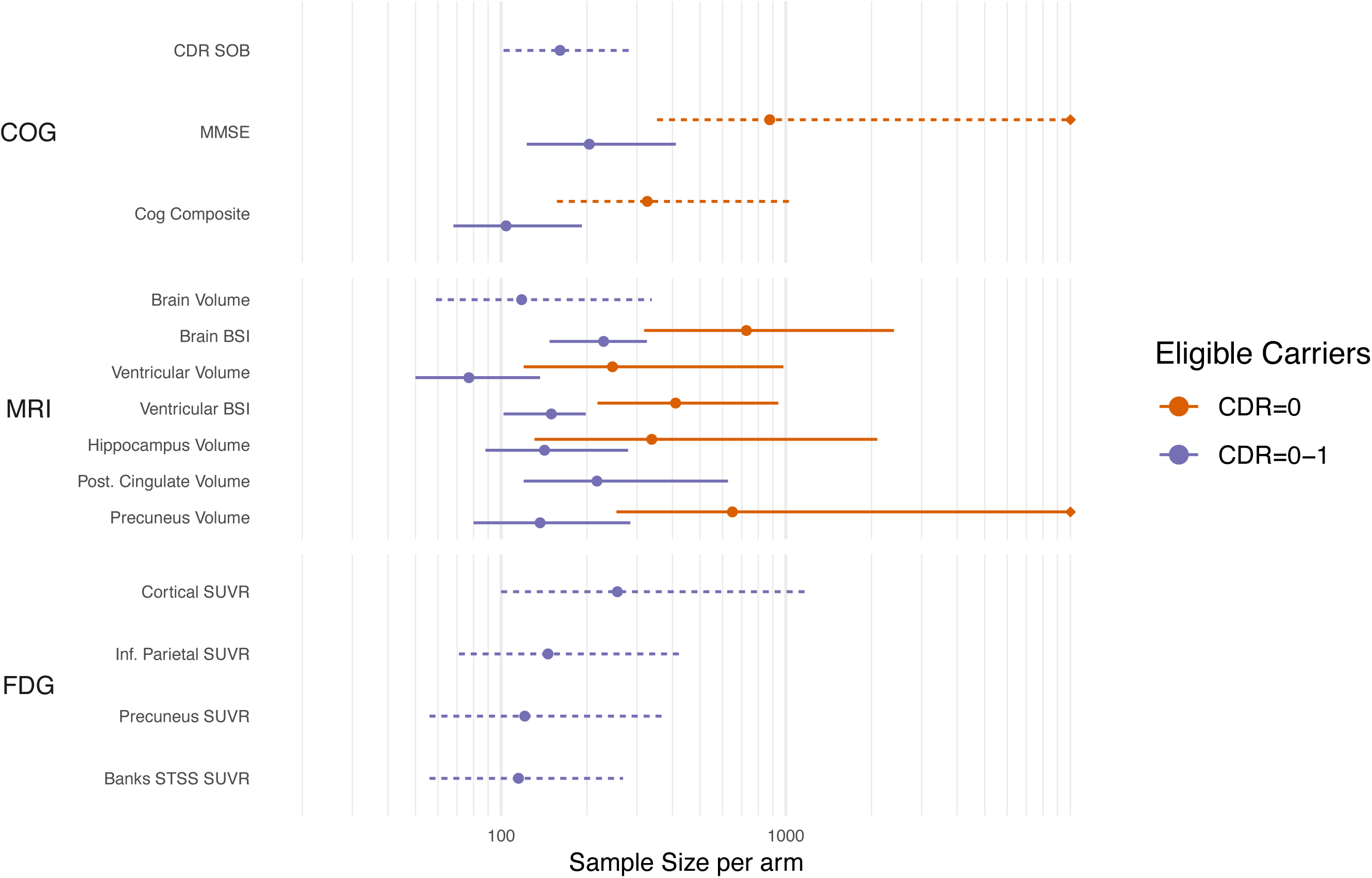
Sample size estimates (with 95% bootstrap confidence intervals) needed to detect a 50% reduction in excess slope (over and above that in non-carriers) over a four-year trial, assuming 40% dropout after four years. Sample size estimates are on a log scale and censored if the higher confidence interval exceeds 10,000. Dashed intervals indicate that the number of bootstrap samples where the model failed to converge was >1%, and thus the confidence intervals should be treated with caution.

The sample size estimates to detect a therapeutic effect of a drug that reduces the final value of the outcome measure by 25% (with respect to the average value in the non-carriers) are shown in Figure 4. Only biomarkers reflecting amyloid or tau pathology were considered in this scenario. For PIB PET measures, sample sizes were similar in both the presymptomaic trial scenario and the one where both unimpaired and mildly impaired participants were eligible.

**Figure 4.**
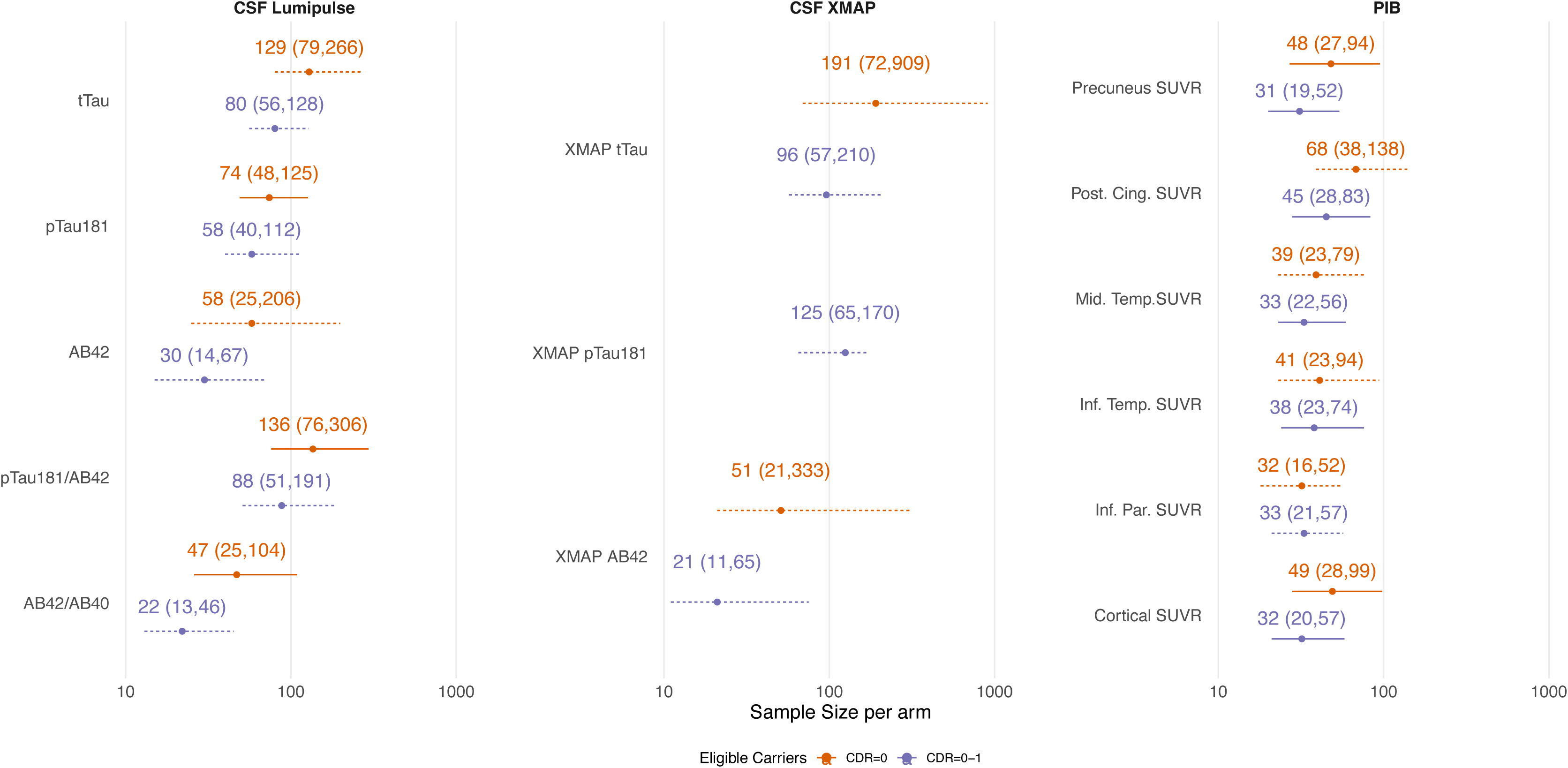
Sample size estimates (with 95% bootstrap confidence intervals) needed to detect a 25% absolute reduction in the untransformed outcome value at the end of the study (relative to the mean level in non-carriers; four-year duration with 40% dropout rate after four years). Sample size estimates are on a log scale. Dashed intervals indicate that the number of bootstrap samples where the model failed to converge was >1%, and thus the confidence intervals should be treated with caution.

## 4. Discussion

Sample size estimates are critical to inform trial designs, particularly in rare diseases like ADAD. To address this, we used data from DIAN-OBS to estimate sample sizes needed to detect clinically relevant treatment effects in individuals with ADAD. We have chosen different levels of target therapeutic effects for biomarkers that measure AD pathology versus those that reflect neurodegeneration. We found that for a trial enrolling only presymptomatic participants, outcome measures of amyloid (via PET and CSF) and tau (via CSF) offered feasible sample sizes for a four-year trial. Sample sizes using MRI and cognitive outcomes in these particpants were much larger. When expanding the eligibility criterion based on global CDR scale to align with the DIAN-TU-001 trial, sample size estimates were much smaller, particularly for markers of neurodegeneration. This is likely due to the differences in rates of accumulation between unimpaired participants and those with mild impairment. These reductions in sample size were far less for PIB PET, likely because of the highly consistent amyloid plaque load and growth in both presymptomatic and early symptomatic stages.

Trials are underway in presymptomatic ADAD. The original DIAN-TU-001 and a study of crenezumab in *PSEN1 E280A* carriers (NCT01998841) have completed. DIAN-TU is recruiting patients for a trial involving lecanemab and the anti-tau agent E2814 (NCT05269394). There is another open label extension of DIAN-TU-001 involving lecanemab (NCT06384573) and a primary prevention study (DIAN-TU-002, NCT03977584). Our results relating to sample sizes for clinical trials in ADAD should be used cautiously when determining sample sizes for future trials in sporadic AD. Some of the outcome measures considered in this paper are more specific to changes observed in ADAD (such as MRI and PET measures in the precuneus). However, many of the outcome measures capture aspects of the disease that are common to both ADAD and sporadic AD (cognitive composite, cortical amyloid accumulation, hippocampal volume, CSF concentrations of Aβ42 and pTau181) and for such outcomes it may not be unreasonable to assume that sample sizes carry across. Further work would be needed to verify this.

### 4.1 Sample sizes to detect a slowing in the rate of neurodegeneration

For four-year trials that include presymptomatic participants (global CDR=0), the sample sizes needed to detect a 50% reduction of slope (relative to non-carriers) in outcomes related to neurodegeneration are likely too high to be feasible in this rare population. The outcomes with the lowest sample size estimates were ventricular volume (246 per arm, 95% CI: [120,980]), the cognitive composite (326 [157, 1074] per arm), and hippocampal volume (338 [131,2096] per arm. No FDG measures had sufficiently different slopes between non-carriers and carriers to include them as a potential outcome measure (see Supplementary Material). Previous results from DIAN-OBS show evidence of increased atrophy and hypometabolism during the presymptomatic stage of the disease. These changes were observed relatively close to expected onset (within five years)[17,37,43], though some ROIs (precuneus, posterior cingulate, banks STS) do show changes as early as 12 years before expected onset.

When including participants with CDR=0-1 inclusive, as was done in DIAN-TU-001, sample size estimates were much smaller. This is likely due to the increased rates of neurodegeneration in individuals who have some evidence of mild impairment (CDR global score of 0.5 and 1) compared to those that are unimpaired. Nine outcome measures (Cognitive: composite; MRI: ventricles, ventricular BSI, whole brain, precuneus and hippocampus; FDG PET: banks STS, inferior parietal, and precuneus) had sample size estimates less than 150 participants per arm. However, caution is warranted when making sample size recommendations for two reasons. First, the upper 95% confidence limits for these estimates (see *4.3 Uncertainty in sample size estimates*; below) are as high as 430 individuals/arm. Second, when a best-performing biomarker is selected from many, the performance of that biomarker is likely to be worse in a new setting due to effects analogous to the well-known phenomenon of regression to the mean.

Ventricular enlargement had some of the lowest sample size requirements across both trial scenarios. Ventricle enlargement can be measured with high precision due to its high-contrast boundaries. The measure is sensitive, but not specific, to pathological atrophy. However, evidence from clinical trials, particularly from anti-amyloid DMTs, indicate that whole brain atrophy and ventricular enlargement may be greater in some treatments compared to placebo[44,45], making its usefulness as an outcome uncertain for this class of DMTs. The biological mechanism driving this phenomenon is unclear. Recently, Belder and colleagues[46] proposed that other mechanisms besides neurodegeneration may result in “amyloid-related pseudo atrophy” (ARPA), and that these excess changes in volume could be caused by reduced inflammation and/or removal of amyloid plaques. It is not yet clear whether similar excess volume changes will occur in other classes of DMTs, such as anti-tau agents.

### 4.2 Sample sizes to detect a reduction of the outcome at the end of the study

From amyloid PET, we obtained sample size estimates of 40-70 participants per arm to detect a 25% reduction in the overall level of amyloid burden (as measured by PIB PET) with 5% statistical significance and 80% power. However, 95% CIs extended up to around 140 participants per arm. Unlike the outcomes related to neurodegeneration, these sample sizes were not substantially reduced when including participants with mild impairment. Sample sizes were comparable when using CSF measures of Aβ42. In both carriers and non-carriers, we found CSF p-tau181 declined over time when using the older XMAP assay, despite substantially increased values in carriers at baseline. This longitudinal decline has been previously observed in symptomatic DIAN-OBS participants[39]. Rates of change in the CSF Lumipulse assay for p-tau181 showed increased rates of change in carriers.

Recent trials have demonstrated that DMTs show large reductions in amyloid burden as measured by PET in individuals with mild AD. In Clarity AD, lecanemab showed evidence of amyloid removal (59 centiloids (77%) decrease from the baseline value of 78 centiloids) [1], while TRAILBLAZER-ALZ 2 reported a reduction of 88 centiloids (87% from the baseline value of 102 centiloids) over 18 months in participants treated with donanemab[2]. These reductions tend to be larger than our proposed target effect and appear to produce a modest cognitive benefit (∼20-30% slowing) in mild AD. However, in the DIAN-TU-001 clinical trial of ADAD, amyloid burden was reduced by 24% over four years in patients treated with gantenerumab compared to the shared placebo arm[12]. While this trial did not show any evidence of cognitive benefit on the primary endpoint, recent results from the open-label extension suggest that asymptomatic participants treated with gantenerumab for the longest duration (∼7-8 years) may experience delays in symptom onset[47]. Combined, these results highlight the complex relationship between amyloid reduction and subsequent slowing in cognitive decline, especially as treatments move earlier in the disease process. We chose the target therapeutic effect of 25% to represent a minimum requirement that would provide a conservative estimate of the sample size required, one that would have a reasonable chance of producing a meaningful clinical benefit in trial scenarios at early stages of the disease process, where the amyloid burden at baseline is much lower in comparison to the recent phase 3 trials. This level of treatment effect may be more plausible for CSF and plasma markers of amyloid and tau, which have shown smaller treatment effects. These sample sizes would provide greater statistical power to detect larger treatment effects. It may be possible to detect larger target treatment effects with fewer participants than we report, but this is not guaranteed. Departures from normality might render the use of ANCOVA inappropriate and hence the basis of our sample size calculations suspect. Variability in the numbers of dropouts (which is reasonable to ignore when sample sizes per arm are large, but not when small) would need to be taken account of in the methodological approach in order to give realistic required sample sizes. For these reasons, we advise not to overrely on predictions using our methodology when these are much below 100 (50 per arm). If larger effects are anticipated, it may be more advisable to carry out trials with shorter durations than four years. These choices will depend on the ability to recruit and retain participants in this rare form of AD, and how effectiveness may vary at shorter durations due to titration regimes that aim to avoid amyloid-related imaging abnormalities (ARIA).

Other markers will be highly relevant to particular therapies. Tau-specific PET tracers are increasingly being included in trials to determine effects on tau burden. Previous longitudinal tau PET studies in ADAD suggest that changes occur very close to expected symptom onset[48,49]. Hence they may be better suited for anti-tau therapies in carriers close to onset.

Recent advances in plasma biomarkers could also serve as potential outcomes. Donanemab reduced plasma concentrations of p-tau217 by about 25% in TRAILBLAZER-ALZ 2[2]. Future work will explore sample sizes required to detect changes in plasma levels of biomarkers such as p-tau217 and neurofilament light.

### 4.3 Uncertainty in sample size estimates

While some point estimates of sample size appear promising, these estimates come with varying degrees of uncertainty, which must be considered when choosing outcome measures for upcoming clinical trials. One way that we can measure the level of uncertainty is to take the ratio between the upper limit on the confidence interval, which could represent a “worst case” scenario for the number of participants needed to sufficiently statistically power a trial, with the lower limit, which could represent a “best case” scenario. For the sample sizes based on presymptomatic trial eligible subjects (CDR=0), the markers producing largest level of uncertainty (as measured by this ratio) were MMSE (ratio above 30), MRI volumetric measures (ratios ranging from 8-40), and CSF Aβ 1-42 (ratio=8.2 for Lumipulse, 15.9 for XMAP). On the other end, PIB PET measures (CI ratio: 3.3-4.1), Lumipulse CSF measures of ptau-181 (ratio=2.6) and Aβ 1-42/1-40 ratio (CI ratio=4.2) and ventricular BSI (CI ratio: 4.3) provided the lowest levels of uncertainty. For the sample sizes based on all trial eligible subjects (CDR=0-1), the markers producing the largest level of uncertainty (as measured by this ratio) were FDG PET markers (ratios between 4.8 to 12), CSF Aβ 1-42 (ratio=4.8 for Lumipulse, 5.9 for XMAP), whole brain volume (ratio=5.7), and posterior cingulate volume (ratio=5.2). On the other end, BSI-based measures of whole brain volume (ratio=2.2) and lateral ventricles (ratio=1.9), cognitive composite(ratio=2.8), and PIB measures (ratio range=2.6-3.2) provided the lowest uncertainty. These measures had the greatest precision in measuring rate of change over time, likely making them more viable for shorter duration trials. The stability of these uncertainty ranges for PIB PET across both trial scenarios is likely due to amyloid accumulation being one of the early observed changes in both sporadic AD and ADAD, with accumulation over time being similar for CDR=0 and CDR=0-1 participants (nearly parallel slopes in Figure 2 and Supplemetal Figure 1).

For many outcomes, particularly CDR-SB, CSF protein levels, FDG-PET and some regional PIB SUVR, there are a high number (> 1%) of bootstrap samples where the LMM failed to fit the data. As the number of bootstrap failures increase, more caution should be given to interpreting the level of uncertainty in the sample size estimates, as it is likely that these missing bootstraps more often represent samples with very high sample size estimates.

Some of the high levels of uncertainty could be attributed to heterogeneity between individuals with ADAD. While ADAD is considered a “pure” form of AD in terms of fewer co-morbidities, heterogeneous patterns of pathology have been observed between mutations in the *PSEN1* and *APP* genes[50], as well as within *PSEN1* mutations[51], which more frequently have atypical phenotypes[6]. Clinical stage is another source of heterogeneity; symptomatic participants have higher rates of atrophy, cognitive decline and hypometabolism compared to presymptomatic carriers, even at mildly symptomatic stages[5,17,35–37]. If there are clear dependencies of endpoints on variables such as CDR®, then one efficient approach would be to stratify at randomisation according to these variables and account for this stratified design in the statistical analysis[34].

This analysis has some limitations. First, there is evidence from the literature [37,39] of non-linear trajectories for some biomarkers in ADAD with respect to EYO, although for the majority of variables, such curvature was not apparent from visual inspection of spaghetti plots of presymptomatic carriers. These non-linearities tend to be observed as individuals approach onset or already have symptoms, so we anticipate that most participants enrolled in presymptomatic trials will be likely to show approximately linear changes over a four-year followup period. Our sample size estimates are theoretically reliant on the assumed linear mixed models being correct. However, small departures from linearity may not materially affect our results. Previous work [52] has shown that in general, these departures have little impact on sample size estimates for clinical trials analysed using mixed models that assume linearity, provided that the treatment effect remains linear and the visit schedule in the observational data and proposed trial are the same. The sensitivity of our findings to assumptions of linearity, particularly when individuals with mild levels of impairment are included, could be further explored in future work. Another limitation is the lack of diversity within the DIAN-OBS study that forms the basis of these sample size estimates. For trials that plan to enroll participants that closely resemble those in DIAN-OBS, our sample size estimates will hopefully be fairly reliable, but this may not be the case in future trials in ADAD if they have a very different demographic makeup, as recent data from studies in sporadic AD suggest that there may be some differences in biomarker findings across different ethnic groups[53–55]. Addressing this lack of diversity is a high priority for DIAN, which is expanding the number of sites globally (Japan, Republic of Korea, Latin America). At existing sites, there are efforts to include ADAD families from different backgrounds into studies. These efforts will increase the diversity of the cohort over time.

Recruiting individuals with such a rare disease can bring challenges. Raising awareness with families around participation in research is essential. DIAN maintains several outreach activities, including a registry (https://dian.wustl.edu/registry/), webinars, and family meetings. While participation in clinical trials can provide a significant burden with potential adverse events, our experience as a local site is that many participants are extremely motivated. Many of these participants begin in DIAN-OBS and then enroll in DIAN-TU. As a result, run-in studies, which have been shown to provide an increase in power[14,33], could be considered. Future work will explore sample size estimates for additional trial designs, such as run-in, common close, and adaptive trials.

## 5. Conclusion

We estimated sample sizes required to detect a clinically meaningful effect within a clinical trial enrolling individuals with ADAD. For presymptomatic trials, detecting a reduction in the absolute level of amyloid burden using PIB PET and CSF of 25% requires sample sizes of 40-70 participants per arm during a four-year trial. Confidence intervals suggest this sample size could need to be as high as around 140 participants per arm. Sample sizes using markers of neurodegeneration tend to be much larger (250-750 participants per arm) to detect a 50% slowing of neurodegeneration over four years. For studies involving both presymptomatic and mildly impaired individuals, volumetric MRI biomarkers require sample sizes spanning 70-230 participants per arm to detect the same effect of slowing. For outcomes related AD pathology of amyloid, sample sizes were comparable in this population. Caution must be exercised when looking at a single estimate of sample size alone, as the uncertainty in this measure can vary significantly, with uncertainty in sample size estimates for FDG PET and CSF tending to be higher than for MRI and PIB PET. Robust sample size estimates are critical to interpret ongoing prevention trials and inform design of upcoming trials in preclinical AD – a stage at which greatest clinical benefit my potentially be achieved.

## Supporting information

DIAN Consortium Author List

## Data Availability

All data produced in the present study are available upon reasonable request to the authors.

https://dian.wustl.edu/our-research/for-investigators/dian-observational-study-investigator-resources/

## Acknowledgements

Data collection and sharing for this project was supported by The Dominantly Inherited Alzheimer’s Network (DIAN, U19AG032438) funded by the National Institute on Aging (NIA), the Alzheimer’s Association (SG-20-690363-DIAN), the German Center for Neurodegenerative Diseases (DZNE), Raul Carrea Institute for Neurological Research (FLENI), Partial support by the Research and Development Grants for Dementia from Japan Agency for Medical Research and Development, AMED, the Korea Dementia Research Project through the Korea Dementia Research Center (KDRC), funded by the Ministry of Health & Welfare and Ministry of Science and ICT, Republic of Korea (HU21C0066) and the Instituto de Salud Carlos III, Spain (grant n° 20/00448 to RSV). This manuscript has been reviewed by DIAN Study investigators for scientific content and consistency of data interpretation with previous DIAN Study publications. We acknowledge the altruism of the participants and their families and the contributions of the DIAN research and support staff at each of the participating sites for their contributions to this study.

## Funding

This work was supported by the National Institute on Aging (DIAN, U19AG032438), the Alzheimer’s Association (SG-20-690363-DIAN), the German Center for Neurodegenerative Diseases (DZNE), Raul Carrea Institute for Neurological Research (FLENI), Research and Development Grants for Dementia from Japan Agency for Medical Research and Development, AMED, the Korea Dementia Research Project through the Korea Dementia Research Center (KDRC), funded by the Ministry of Health & Welfare and Ministry of Science and ICT, Republic of Korea (HU21C0066), the Instituto de Salud Carlos III, Spain (grant n° 20/00448 to RSV), the Alzheimer’s Society (AS-PG-15-025), the UK Dementia Research Institute which receives its funding from DRI Ltd, funded by the UK Medical Research Council, Alzheimer’s Society and Alzheimer’s Research UK, the NIHR University College London Hospitals Biomedical Research Centre, and the UKRI Medical Research Council (Skills development fellowship MR/P014372/1).

## Declaration of Generative AI and AI-assisted technologies in the writing process

I confirm that no generative AI or AI-assisted technology was used in the writing process.

## Notes

### Competing Interest Statement

DMC reports consultancy fees for Perceptive Imaging.
JJLG is supported by NIH-NIA (K01AG073526), the Alzheimers Association (AARFD-21-851415, SG-20-690363), the Michael J. Fox Foundation (MJFF-020770), the Foundation for Barnes Jewish Hospital and the McDonnell Academy.
EMD received support from the National Institute on Aging, an anonymous organization, the GHR Foundation, the DIAN-TU Pharma Consortium, Eli Lilly, and F Hoffmann La-Roche; has received speaking fees from Eisai and Eli Lilly; and is on the data safety and monitoring board and advisory boards of Eli Lilly, Alector, and Alzamend.
GD reports no competing interests directly relevant to this work. His research is supported by NIH (K23AG064029, U01AG057195, U01NS120901, U19AG032438). He serves as a consultant for Parabon Nanolabs Inc and as a Topic Editor (Dementia) for DynaMed (EBSCO). He is the co-Project PI for a clinical trial in anti-NMDAR encephalitis, which receives support from NINDS (U01NS120901) and Amgen Pharmaceuticals; and a consultant for Arialys Therapeutics. He has developed educational materials for Continuing Education Inc and Ionis Pharmaceutical. He owns stock in ANI pharmaceuticals. Dr. Day's institution has received support from Eli Lilly for development and participation in an educational event promoting early diagnosis of symptomatic Alzheimer disease, and in-kind contributions of radiotracer precursors for tau-PET neuroimaging in studies of memory and aging (via Avid Radiopharmaceuticals, a wholly owned subsidiary of Eli Lilly).
PRS receives funding from the National Health and Medical Research Council (Australia) grants 1176716 and 2022057 and the Medical Research Future Fund (Australia) grants 1200428 and 1200428. He is a director (unpaid) of the Australian Dementia Network Ltd.
RJB, Professor of Neurology at Washington University School of Medicine (WUSM) receives lab research funding from the National Institutes of Health, Alzheimers Association, BrightFocus Foundation, Rainwater Foundation, Association for Frontotemporal Degeneration FTD Biomarkers Initiative, Tau Consortium, Novartis, Centene Corporation, Association for Frontotemporal Degeneration, the Cure Alzheimers Fund, Coins for Alzheimers Research Trust Fund, The Foundation for Barnes-Jewish Hospital, Good Ventures Foundation, DIAN-TU Pharma Consortium, Centene Corporation, Tau SILK Consortium (AbbVie, Biogen, Eli Lilly and Company and an anonymous organization), the NfL Consortium (AbbVie, Biogen, Bristol Meyers Squibb, Hoffman La Roche, and an anonymous organization). RJB has received honoraria as a speaker/consultant/advisory board member from Eisai, F. Hoffman-LaRoche, Janssen, Biogen; and reimbursement of travel expenses from Korean Dementia Association, American Neurological Association, Fondazione Prada, Weill Cornell Medical College, Harvard University, CTAD, FBRI, Beeson Foundation, Adler, Alzheimers Association Roundtable, Duke Margolis Roundtable, Bright Focus Foundation, Tau Consortium Investigators, NAPA Advisory Council on Alzheimers Research. RJB serves as principal investigator of the DIAN-TU, which is supported by the Alzheimers Association, GHR Foundation, an anonymous organization and the DIAN-TU Pharma Consortium (Active: Biogen, Eisai, Eli Lilly and Company/Avid Radiopharmaceuticals, F. Hoffman-La Roche/Genentech, and Janssen. Previous: Abbvie, Amgen, AstraZeneca, Forum, Mithridion, Novartis, Pfizer, Sanofi, and United Neuroscience). The DIAN-TU-001 Clinical Trial is supported by Pharmaceutical Partners Eli Lilly and Company, F. Hoffman-La Roche and Janssen, the Alzheimers Association, NIH U01AG042791, NIH U01AG42791-S1 (FNIH and Accelerating Medicines Partnership), NIH R01AG046179, NIH R56AG053267, NIH R01AG053267, NIH U01AG059798, NIH R01AG068319, Avid Radiopharmaceuticals, GHR Foundation, and an anonymous organization. In-kind support has been received from CogState, Cerveau, Signant Health and Eisai Corporation. RJB is a co-founder of C2N Diagnostics and receives income from C2N Diagnostics for serving on the scientific advisory board. Washington University (WU) has equity ownership interest in C2N Diagnostics. C2N Diagnostics will be analyzing samples from the Knight Family DIAN-TU-001 trial of E2814 for primary, secondary, and exploratory endpoints. Should the DIAN-TU trials impact the value of C2N Diagnostics, WU and RJB could directly benefit.
NCF reports consulting fees from Biogen, Eisai, Ionis, Lilly, Roche/Genentech, and Siemens paid to UCL; he has served on a Data Safety Monitoring Board for Biogen; he acknowledges grant support from the Alzheimers Society, Alzheimers Research UK, Rosetrees Trust, the Sigrid Rausing Trust, the UK Dementia Research Institute and the UK NIHR UCLH Biomedical Research Centre.
Johannes Levin reports speaker fees from Bayer Vital, Biogen, EISAI, TEVA, Zambon, Esteve, Merck and Roche, consulting fees from Axon Neuroscience, EISAI and Biogen, author fees from Thieme medical publishers and W. Kohlhammer GmbH medical publishers and is inventor in a patent "Oral Phenylbutyrate for Treatment of Human 4-Repeat Tauopathies" (EP 23 156 122.6) filed by LMU Munich. In addition, he reports compensation for serving as chief medical officer for MODAG GmbH, is beneficiary of the phantom share program of MODAG GmbH and is inventor in a patent "Pharmaceutical Composition and Methods of Use" (EP 22 159 408.8) filed by MODAG GmbH, all activities outside the submitted work.
All other authors have no competing interests to disclose.

### Author Declarations

The Institutional Review Board (IRB) of Washington University School of Medicine in St. Louis approved the study with the IRB ID# 201106339, and research was performed in accordance with the approved protocols The Ethics Committee of University College London (South Central-Berkshire ethics committe) gave ethics approval for this work under reference 09/HA0505/73.

### Summary of Updates

This revision addresses various elements picked up in peer review and places a focus on the presymptomatic trial design with the DIAN-TU-001 eligibility crieteria as a benchmark

## References

[1] van Dyck CH, Swanson CJ, Aisen P, Bateman RJ, Chen C, Gee M, et al. Lecanemab in Early Alzheimer’s Disease. New England Journal of Medicine 2022:1–13. 10.1056/NEJMoa2212948.

[2] Sims JR, Zimmer JA, Evans CD, Lu M, Ardayfio P, Sparks J, et al. Donanemab in Early Symptomatic Alzheimer Disease. JAMA 2023;330:512. 10.1001/jama.2023.13239.

[3] Budd Haeberlein S, Aisen PS, Barkhof F, Chalkias S, Chen T, Cohen S, et al. Two Randomized Phase 3 Studies of Aducanumab in Early Alzheimer’s Disease. J Prev Alzheimers Dis 2022;9:197–210. 10.14283/jpad.2022.30.

[4] Villemagne VL, Burnham S, Bourgeat P, Brown B, Ellis KA, Salvado O, et al. Amyloid β deposition, neurodegeneration, and cognitive decline in sporadic Alzheimer’s disease: a prospective cohort study. Lancet Neurology 2013;12:357–67. 10.1016/S1474-4422(13)70044-9.

[5] Bateman RJ, Xiong C, Benzinger TLS, Fagan AM, Goate A, Fox NC, et al. Clinical and Biomarker Changes in Dominantly Inherited Alzheimer’s Disease. New England Journal of Medicine 2012;367:795–804. 10.1056/NEJMoa1202753.

[6] Ryan NS, Nicholas JM, Weston PSJ, Liang Y, Lashley T, Guerreiro R, et al. Clinical phenotype and genetic associations in autosomal dominant familial Alzheimer’s disease: a case series. Lancet Neurol 2016;15:1326–35. 10.1016/S1474-4422(16)30193-4.

[7] Ryman DC, Acosta-Baena N, Aisen PS, Bird T, Danek A, Fox NC, et al. Symptom onset in autosomal dominant Alzheimer disease: A systematic review and meta-analysis. Neurology 2014;83:253–60. 10.1212/WNL.0000000000000596.

[8] Sperling R, Mormino E, Johnson K. The Evolution of Preclinical Alzheimer’s Disease: Implications for Prevention Trials. Neuron 2014;84:608–22. 10.1016/j.neuron.2014.10.038.

[9] Insel PS, Weiner M, Scott MacKin R, Mormino E, Lim YY, Stomrud E, et al. Determining clinically meaningful decline in preclinical Alzheimer disease. Neurology 2019;93:E322–33. 10.1212/WNL.0000000000007831.

[10] US Food and Drug Administration. Early Alzheimer’s Disease: Developing Drugs for Treatment; Draft Guidance for Industry. 2018.

[11] Rafii MS, Sperling RA, Donohue MC, Zhou J, Roberts C, Irizarry MC, et al. The AHEAD 3-45 Study: Design of a prevention trial for Alzheimer’s disease. Alzheimer’s & Dementia 2023;19:1227–33. 10.1002/ALZ.12748.

[12] Salloway S, Farlow M, McDade E, Clifford DB, Wang G, Llibre-Guerra JJ, et al. A trial of gantenerumab or solanezumab in dominantly inherited Alzheimer’s disease. Nat Med 2021;27:1187–96. 10.1038/s41591-021-01369-8.

[13] Moulder KL, Snider BJ, Mills SL, Buckles VD, Santacruz AM, Bateman RJ, et al. Dominantly Inherited Alzheimer Network: facilitating research and clinical trials. Alzheimers Res Ther 2013;5:48. 10.1186/alzrt213.

[14] Wang G, Aschenbrenner AJ, Li Y, McDade E, Liu L, Benzinger TLS, et al. Two-period linear mixed effects models to analyze clinical trials with run-in data when the primary outcome is continuous: Applications to Alzheimer’s disease. Alzheimer’s and Dementia: Translational Research and Clinical Interventions 2019;5:450–7. 10.1016/j.trci.2019.07.007.

[15] Morris JC, Aisen PS, Bateman RJ, Benzinger TLS, Cairns NJ, Fagan AM, et al. Developing an international network for Alzheimer research: The Dominantly Inherited Alzheimer Network. Clin Investig (Lond) 2012;2:975–84. 10.4155/cli.12.93.

[16] Morris JC. The Clinical Dementia Rating (CDR): current version and scoring rules. Neurology 1993;43:2412–4.

[17] Benzinger TLS, Blazey T, Jack Jr. CR, Koeppe RA, Su Y, Xiong C, et al. Regional variability of imaging biomarkers in autosomal dominant Alzheimer’s disease. Proc Natl Acad Sci U S A 2013;110:E4502–9. 10.1073/pnas.1317918110.

[18] Cardoso MJ, Modat M, Wolz R, Melbourne A, Cash DM, Rueckert D, et al. Geodesic Information Flows: Spatially-Variant Graphs and Their Application to Segmentation and Fusion. IEEE Trans Med Imaging 2015;34:1976–88. 10.1109/TMI.2015.2418298.

[19] Malone IB, Leung KK, Clegg S, Barnes J, Whitwell JL, Ashburner J, et al. Accurate automatic estimation of total intracranial volume: A nuisance variable with less nuisance. Neuroimage 2015;104:366–72. 10.1016/j.neuroimage.2014.09.034.

[20] Leung KK, Clarkson MJ, Bartlett JW, Clegg S, Jack Jr. CR, Weiner MW, et al. Robust atrophy rate measurement in Alzheimer’s disease using multi-site serial MRI: Tissue-specific intensity normalization and parameter selection. Neuroimage 2010;50:516–23. 10.1016/j.neuroimage.2009.12.059.

[21] Prados F, Cardoso MJ, Leung KK, Cash DM, Modat M, Fox NC, et al. Measuring brain atrophy with a generalized formulation of the boundary shift integral. Neurobiol Aging 2015;36:S81--S90.

[22] Freeborough PA, Fox NC. The boundary shift integral: an accurate and robust measure of cerebral volume changes from registered repeat MRI. IEEE Trans Med Imaging 1997;16:623–9. 10.1109/42.640753.

[23] Su Y, Angelo GMD, Vlassenko AG, Zhou G, Snyder AZ, Marcus DS, et al. Quantitative Analysis of PiB-PET with FreeSurfer ROIs 2013;8. 10.1371/journal.pone.0073377.

[24] Mann DMA, Iwatsubo T, Ihara Y, Cairns NJ, Lantos PL, Bogdanovic N, et al. Predominant deposition of amyloid-β42(43) in plaques in cases of Alzheimer’s disease and hereditary cerebral hemorrhage associated with mutations in the amyloid precursor protein gene. American Journal of Pathology 1996;148:1257–66.

[25] Ghisays V, Lopera F, Goradia DD, Protas HD, Malek-Ahmadi MH, Chen Y, et al. PET evidence of preclinical cerebellar amyloid plaque deposition in autosomal dominant Alzheimer’s disease-causing Presenilin-1 E280A mutation carriers. Neuroimage Clin 2021;31:102749. 10.1016/J.NICL.2021.102749.

[26] Rousset OG, Ma Y, Evans AC. Correction for partial volume effects in PET: Principle and validation. Journal of Nuclear Medicine 1998;39:904–11.

[27] Su Y, Blazey TM, Snyder AZ, Raichle ME, Marcus DS, Ances BM, et al. NeuroImage Partial volume correction in quantitative amyloid imaging 2015;c:55–64. 10.1016/j.neuroimage.2014.11.058.

[28] Fagan AM, Xiong C, Jasielec MS, Bateman RJ, Goate AM, Benzinger TLS, et al. Longitudinal Change in CSF Biomarkers in Autosomal-Dominant Alzheimer’s Disease. Sci Transl Med 2014;6:226ra30–226ra30. 10.1126/scitranslmed.3007901.

[29] Karran E, Mercken M, Strooper B De. The amyloid cascade hypothesis for Alzheimer’s disease: An appraisal for the development of therapeutics. Nat Rev Drug Discov 2011;10:698–712. 10.1038/nrd3505.

[30] Mintun MA, Lo AC, Duggan Evans C, Wessels AM, Ardayfio PA, Andersen SW, et al. Donanemab in Early Alzheimer’s Disease. New England Journal of Medicine 2021;384:1691–704. 10.1056/NEJMoa2100708.

[31] Toyn JH, Ahlijanian MK. Interpreting Alzheimer’s disease clinical trials in light of the effects on amyloid-β. Alzheimers Res Ther 2014;6. 10.1186/alzrt244.

[32] Salloway S, Farlow M, McDade E, Clifford DB, Wang G, Llibre-Guerra JJ, et al. A trial of gantenerumab or solanezumab in dominantly inherited Alzheimer’s disease. Nat Med 2021;27:1187–96. 10.1038/s41591-021-01369-8.

[33] Frost C, Kenward MG, Fox NC. Optimizing the design of clinical trials where the outcome is a rate . Can estimating a baseline rate in a run-in period increase efficiency? Stat Med 2008;27:3717–31. 10.1002/sim.

[34] Frost C, Mulick A, Scahill RI, Owen G, Aylward E, Leavitt BR, et al. Design optimization for clinical trials in early-stage manifest Huntington’s disease. Movement Disorders 2017;32:1610–9. 10.1002/mds.27122.

[35] Cash DM, Ridgway GR, Liang Y, Ryan NS, Kinnunen KM, Yeatman T, et al. The pattern of atrophy in familial Alzheimer disease: volumetric MRI results from the DIAN study. Neurology 2013;81:1425–33. 10.1212/WNL.0b013e3182a841c6.

[36] Kinnunen KM, Cash DM, Poole T, Frost C, Benzinger TLSS, Ahsan RL, et al. Presymptomatic atrophy in autosomal dominant Alzheimer’s disease: A serial magnetic resonance imaging study. Alzheimers Dement 2018;14:43–53. 10.1016/j.jalz.2017.06.2268.

[37] Gordon BA, Blazey TM, Su Y, Hari-Raj A, Dincer A, Flores S, et al. Spatial patterns of neuroimaging biomarker change in individuals from families with autosomal dominant Alzheimer’s disease: a longitudinal study. Lancet Neurol 2018;17:241–50. 10.1016/S1474-4422(18)30028-0.

[38] Wang G, Berry S, Xiong C, Hassenstab J, Quintana M, McDade EM, et al. A novel cognitive disease progression model for clinical trials in autosomal-dominant Alzheimer’s disease. Stat Med 2018;37:3047–55. 10.1002/sim.7811.

[39] McDade E, Wang G, Gordon BA, Hassenstab J, Benzinger TLS, Buckles V, et al. Longitudinal cognitive and biomarker changes in dominantly inherited Alzheimer disease. Neurology 2018:10.1212/WNL.0000000000006277. 10.1212/WNL.0000000000006277.

[40] Bateman RJ, Benzinger TL, Berry S, Clifford DB, Duggan C, Fagan AM, et al. The DIAN-TU Next Generation Alzheimer’s prevention trial: Adaptive design and disease progression model. Alzheimer’s & Dementia 2017;13:8–19. 10.1016/j.jalz.2016.07.005.

[41] Sperling RA, Donohue MC, Raman R, Rafii MS, Johnson K, Masters CL, et al. Trial of Solanezumab in Preclinical Alzheimer’s Disease. New England Journal of Medicine 2023;12:1096–107. 10.1056/NEJMOA2305032/SUPPL_FILE/NEJMOA2305032_DATA-SHARING.PDF.

[42] Nash S, Morgan KE, Frost C, Mulick A. Power and sample-size calculations for trials that compare slopes over time: Introducing the slopepower command: Stata J 2021;21:575–601. 10.1177/1536867X211045512.

[43] Kinnunen KM, Cash DM, Poole T, Frost C, Benzinger TLS, Ahsan RL, et al. Presymptomatic atrophy in autosomal dominant Alzheimer’s disease: A serial magnetic resonance imaging study. Alzheimers Dement 2018;14:43–53. 10.1016/j.jalz.2017.06.2268.

[44] Cash DM, Rohrer JD, Ryan NS, Ourselin S, Fox NC. Imaging endpoints for clinical trials in Alzheimer’s disease. Alzheimers Res Ther 2014;6:87. 10.1186/s13195-014-0087-9.

[45] Schwarz AJ. The Use, Standardization, and Interpretation of Brain Imaging Data in Clinical Trials of Neurodegenerative Disorders. Neurotherapeutics 2021;18:686–708. 10.1007/s13311-021-01027-4.

[46] Belder CRS, Boche D, Nicoll JAR, Jaunmuktane Z, Zetterberg H, Schott JM, et al. Brain volume change following anti-amyloid β immunotherapy for Alzheimer’s disease: amyloid-removal-related pseudo-atrophy. Lancet Neurol 2024;23:1025–34. 10.1016/S1474-4422(24)00335-1.

[47] Bateman RJ, Li Y, McDade E, Llibre Guerra JJ, Clifford D, Atri A, et al. Amyloid Reduction and Dementia Progression in Dominantly Inherited Alzheimer’s Diseaseafter Long-Term Gantenerumab Treatment: Results from the Dian-Tu Trial n.d. 10.2139/SSRN.4906344.

[48] O’Connor A, Cash DM, Poole T, Markiewicz PJ, Fraser MR, Malone IB, et al. Tau accumulation in autosomal dominant Alzheimer’s disease: a longitudinal [18F]flortaucipir study. Alzheimers Res Ther 2023;15:1–11. 10.1186/S13195-023-01234-5/FIGURES/3.

[49] Gordon BA, Blazey TM, Christensen J, Dincer A, Flores S, Keefe S, et al. Tau PET in autosomal dominant Alzheimer’s disease: relationship with cognition, dementia and other biomarkers. Brain 2019;142:1063–76. 10.1093/brain/awz019.

[50] Scahill RI, Ridgway GR, Bartlett JW, Barnes J, Ryan NS, Mead S, et al. Genetic Influences on Atrophy Patterns in Familial {A}lzheimer’s Disease: A Comparison of {APP} and {PSEN1} Mutations. J Alzheimers Dis 2013. 10.3233/JAD-121255.

[51] Chhatwal JP, Schultz SA, McDade E, Schultz AP, Liu L, Hanseeuw BJ, et al. Variant-dependent heterogeneity in amyloid β burden in autosomal dominant Alzheimer’s disease: cross-sectional and longitudinal analyses of an observational study. Lancet Neurol 2022;21:140–52. 10.1016/S1474-4422(21)00375-6.

[52] Morgan KE, White IR, Frost C. How important is the linearity assumption in a sample size calculation for a randomised controlled trial where treatment is anticipated to affect a rate of change? BMC Med Res Methodol 2023;23:1–19. 10.1186/S12874-023-02093-2/TABLES/1.

[53] Gottesman RF, Schneider ALC, Zhou Y, Chen X, Green E, Gupta N, et al. The ARIC-PET amyloid imaging study: Brain amyloid differences by age, race, sex, and APOE. Neurology 2016;87:473–80. 10.1212/WNL.0000000000002914.

[54] Wilkins CH, Windon CC, Dilworth-Anderson P, Romanoff J, Gatsonis C, Hanna L, et al. Racial and Ethnic Differences in Amyloid PET Positivity in Individuals With Mild Cognitive Impairment or Dementia: A Secondary Analysis of the Imaging Dementia–Evidence for Amyloid Scanning (IDEAS) Cohort Study. JAMA Neurol 2022;79:1139–47. 10.1001/JAMANEUROL.2022.3157.

[55] Deters KD, Napolioni V, Sperling RA, Greicius MD, Mayeux R, Hohman T, et al. Amyloid PET Imaging in Self-Identified Non-Hispanic Black Participants of the Anti-Amyloid in Asymptomatic Alzheimer’s Disease (A4) Study. Neurology 2021;96:E1491–500. 10.1212/WNL.0000000000011599.

