## Supplemental Material for "Sample size estimates for biomarker-based outcome measures in clinical trials in autosomal dominant Alzheimer’s disease"

**Statistical appendix**

1. Overview

We assess sample size estimates for various trial designs using a two-stage approach also adopted in previous work[1,2], and here we describe this process in generality. In summary: in the first stage we fit linear mixed models (LMM) to observational repeated measures data from carriers and non-carriers in the DIAN-OBS study. From the models fitted to carriers, we obtain estimates of the key parameters describing variability over time: these being the parameters that inform sample size calculations for planned clinical trials. We also estimate (using the models for carriers and for non-carriers) plausible target therapeutic effects. In the second stage we use the estimated variability parameters and target treatment effects to estimate the sample size requirement for a postulated trial. The postulated trial can potentially include designs with multiple follow-up visits, but here we restrict ourselves to simple designs with a baseline visit and a single follow-up visit. Both stages are described in detail in Section 2 below.

The basic two-stage approach can be extended to provide confidence intervals for sample sizes. The method to do this is described in Section 3. This is followed (in Section 4) by computational details.

2. The two-stage approach to sample size estimation

*Stage 1a: Quantifying variability from models for repeated measures*

i) *All Cognitive, CSF, PIB and FDG variables and all MRI variables other than those based on the boundary shift integral*

For all outcome measures, except those based on the boundary shift integral, standard random slopes models were fitted separately to carriers and non-carriers. These LMMs have the form:

$y_{ij}=(\beta_{0}{+ b}_{0i})+(\beta_{1}{+ b}_{1i})t_{ij}+e_{ij}$ with $\left( \begin{matrix} b_{0i} \\ b_{1i} \end{matrix} \right)\sim N\left( 0,\left( \begin{matrix} \sigma_{0}^{2} & \sigma_{01} \\ \sigma_{01} & \sigma_{1}^{2} \end{matrix} \right) \right)$ and $e_{ij}\sim N\left( 0,\sigma_{e}^{2} \right)$

(1)

where $y_{ij}$ is the value of the outcome variable for the $i^{th}$ person at the $j^{th}$ visit and $t_{ij}$ is the time of that visit relative to the start of follow up, with the first visit for each person occurring at time $t_{i0}=0$. In the model for non-carriers the random effect for slope was omitted (i.e. $\sigma_{01}$ and $\sigma_{1}^{2}$ were set to zero) since variation in change over time between non-carriers is generally very small, and leaving such a term in the model in such a case can lead to convergence problems.

For the MRI, FDG, CSF and PIB variables (but not the cognitive variables) outcomes were log-transformed in order to make changes over time more approximately linear. When log-transformed (and after multiplying by 100), a slope essentially represents change on a percentage scale with respect to baseline. The models for volumetric MRI measures were extended to include fixed effects for total intracranial volume (TIV) and its interaction with time. TIV was centered by subtracting the mean of TIV in carriers from each TIV measurement, such that the interpretation of $\beta_{1}$ from the model represented by equation (1) is the slope over time in a participant with the mean TIV observed in those with ADAD.

ii) *MRI variables based on the boundary shift integral*

For “direct” measurements of change between two visits obtained from the boundary shift integral, a different LMM was used[3,4]:

$c_{ijk}=\left( \beta_{1}{+ b}_{1i} \right)\left( t_{ik}-t_{ij} \right){- u}_{ij}+u_{ik}+w_{ijk}$with $b_{1i}\sim N\left( 0,\sigma_{b1}^{2} \right), u_{ij}\sim N\left( 0,\sigma_{u}^{2} \right)$

and$w_{ijk}\sim N(0,\sigma_{w}^{2})$	(2)

where$\text{ }c_{ijk}$is the measured change in the outcome variable for participant *i* between visits *j* and *k*, $u_{ij}$ and $\text{ }u_{ik}$are subject-specific random visit effects on the direct measurement of change, and $w_{ijk}$ is the unexplained residual variability. Other notation is as above.

Prior to fitting the model boundary shift integral derived changes were converted to change as a proportion of baseline, defined as *p*, by dividing by brain volume at the start of the relevant time interval. For consistency with the log-transformation approach in *i)* these proportions were transformed using 100*$\log_{e}\left( 1+p \right)$ (for further information around this transformation, please see [3].

*Stage 1b: Target therapeutic effects*

i) *Cognitive, MRI and FDG PET variables*

In considering target therapeutic effects, a useful starting point is to understand what a 100% effective (*i.e.* completely successful) treatment might achieve. For outcomes reflecting neurodegeneration (cognitive, MRI and FDG variables in our study) a 100% effective treatment effect might be expected to reduce the mean *rate of change* in that variable to that in non-carriers. For such outcomes (irrespective of whether analysed using equation (1) or (2)), we consider an appropriate target therapeutic effect to be a $p\times100$% reduction in the excess mean rate of change (over and above that seen in non-carriers) (see left panel of Figure 1, where *p* is taken to be 0.5). Extending the notation in equations (1) and (2) such that $\beta_{1C}$ and $\beta_{1NC}$represent the mean rates of change in carriers and non-carriers respectively, the trial should use a target therapeutic effect that reduces the rate of change from $\beta_{1C}$ in the placebo arm to $\beta_{1C}-p\left( \beta_{1C}-\beta_{1NC} \right)$in the intervention arm. The target treatment effect for these variables can be left as the difference between the slopes in the two groups, $-p\left( \beta_{1C}-\beta_{1NC} \right)$, or converted to the difference between the mean levels in the two groups at the end of the follow-up period of the trial (i.e. at time $t$):

$$\begin{aligned} d=-pt\left( \beta_{1C}-\beta_{1NC} \right)\#\left( 3 \right) \end{aligned}$$

For the model of direct measures of change represented by equation (2) this quantity corresponds to the difference in mean change between baseline and time $t$.

For the MRI and FDG variables (but not the cognitive variables) changes over time are more approximately linear after log-transformation. When log-transformed, a slope essentially represents change on a percentage scale with respect to baseline and so a $p\times100$% reduction in the excess mean rate of change relates to rates of change that are themselves on a percentage scale.

ii) *CSF and PIB variables*

For outcomes reflective of amyloid and/or tau burden (CSF and PIB variables), a 100% effective treatment effect might be expected to reduce the average *level* of that variable to that in non-carriers. For such outcomes we consider an appropriate target therapeutic effect to be $p\times100$% of the difference between the averages in carriers and non-carriers at the end of follow-up (see right panel of Figure 1, where *p* is taken to be 0.25).

As for the MRI and FDG variables above, log-transforms were used for the CSF and PIB variables. However, target treatment effects were defined on the original untransformed scale, and considered geometric means to represent average levels. Extending the notation in equation (1) such that $\beta_{0C}$ and $\beta_{0NC}$ represent the mean baseline levels of the log-transformed outcome in carriers and non-carriers respectively and *t* is the length of follow-up in the postulated trial, the target therapeutic effect reduces the geometric mean level on the untransformed scale at the end of follow-up from $e^{\beta_{0C}+\beta_{1C}t}$ in the placebo arm to $(1-p)e^{\beta_{0C}+\beta_{1C}t}+pe^{\beta_{0NC}+\beta_{1NC}t}$ in the intervention arm. When the outcome is log-transformed this equates to a reduction from an (arithmetic) mean of $\beta_{0C}+\beta_{1C}t$ in the placebo arm to one of $\text{log}\left( (1-p)e^{\beta_{0C}+\beta_{1C}t}+pe^{\beta_{0NC}+\beta_{1NC}t} \right)$ in the intervention arm. The target treatment effect for these variables is the difference between the mean log-transformed levels at time $t$:

$$\begin{aligned} d=-\left( \beta_{0C}+\beta_{1C}t-\text{log}\left( (1-p)e^{\beta_{0C}+\beta_{1C}t}+pe^{\beta_{0NC}+\beta_{1NC}t} \right) \right)\#\left( 4 \right) \end{aligned}$$

We also make the commonly adopted assumption in all scenarios that variability will be unaltered by treatment: such that variability in both arms will mimic that seen in the carrier group in the observational data.

*Stage 2: Sample sizes for clinical trial designs*

A number of authors including Dawson and Frost and colleagues[2,5] have shown how required sample sizes for a particular clinical trial design that is to be analysed using an LMM can be computed provided that there are postulated values for the parameters in the LMM, and a target treatment effect defined as a difference in slopes in the two treatment groups. One requirement is that transformations are used in a consistent fashion: for example, where the observational data are log-transformed then a log-transformation must also be used in the analysis of the trial.

i) *All trials with a baseline and single follow-up measure of a Cognitive, CSF, PIB, FDG or MRI variable*

While the general approach for any LMM can be used here, for a simple planned design with just a single baseline and follow-up measure at time *t*, the simplest (and essentially equivalent) appropriate approach is to use an analysis of covariance (ANCOVA) model. Frison and Pocock[6] give a formula for the sample size in each arm of a trial for the ANCOVA treatment effect estimator:

$$\begin{aligned} N=\frac{2\left( \sigma_{F}^{2}-\frac{\sigma_{BF}^{2}}{\sigma_{B}^{2}} \right)}{d^{2}}\left( 1.96+0.842 \right)^{2}\#\left( 5 \right) \end{aligned}$$

for a power of 80% and a Type I error of 5%, where $d$ is the target treatment effect defined as a difference in means between the two groups at time $t$ (equation (3) for cognitive, MRI and FDG variables, equation (4) for CSF and PIB variables), $\sigma_{B}^{2}$ is the variance of the baseline measure, $\sigma_{F}^{2}$ is the variance of the follow-up measure at time $t$, and $\sigma_{BF}$ is the covariance between the follow-up and baseline measures. These can be estimated from the repeated measures model specified in equation (1). Specifically, the implied variance at time *t,* $\sigma_{F}^{2},$ is $\sigma_{0}^{2}+ 2t\sigma_{01}+ {t^{2}\sigma}_{1}^{2}+\sigma_{e}^{2}$, the implied variance at baseline, $\sigma_{B}^{2}$, is $\sigma_{0}^{2}+\sigma_{e}^{2}$, and the implied covariance with the baseline measure, $\sigma_{BF}$, is$\text{ }\sigma_{0}^{2}+ t\sigma_{01}$.

As explained in Frost et. al.[1], for designs where it is planned to additionally adjust for a covariate such as TIV, the variances of the baseline and follow-up measures conditional on the covariate (and the covariance between the follow-up and baseline measures conditional on the covariate) should be used. Estimates of these conditional (co)variances can be obtained by including the centred covariate and its interaction with time in the models specified in equation (1). Assuming that covariates are perfectly balanced by randomisation arm, the estimates of the covariate effects are orthogonal to those for the treatment effect and hence neither the covariate effect estimates, nor their variances and covariances, need be used in the formulae for the calculation of sample-sizes (equation (5)) provided the covariate and its interaction with time are added to the model specified in equation (1). Centring the covariate ensures that the target therapeutic effect is specified for a person with mean levels of the covariate.

ii) *Trials with a single “direct” measure of change*

For a planned design with just a single “direct” measure of change at time *t*, the simplest appropriate approach is to use an unpaired t test. The appropriate formula for the sample size in each arm of the trial is:

$$\begin{aligned} N=\frac{2\sigma_{C}^{2}}{d^{2}}\left( 1.96+0.842 \right)^{2}\#\left( 6 \right) \end{aligned}$$

for a power of 80% and a Type I error of 5%, where $d$ is the target treatment effect defined as a difference in mean changes between baseline and time $t$ between the two groups (equation (3)) and $\sigma_{C}^{2}$ is the variance of the change over time $t$. These can be estimated from the repeated measures model specified in equation (2). Specifically, the implied variance of the change over time *t,* $\sigma_{C}^{2},$ is ${t^{2}\sigma_{b1}^{2}+2\sigma}_{u}^{2}+\sigma_{w}^{2}$.

iii) *Allowing for dropout*

Once the sample size is calculated using equation (5) or (6), any dropout at the follow-up time point can be accounted for by inflating the value of $N$. For example, for $s\times100\%$ dropout at time $t$, the required sample size $N$ can be calculated from the sample size with no dropout, $N_{complete}$, as follows:

$$\begin{aligned} N=\frac{N_{complete}}{1-s}\#\left( 7 \right) \end{aligned}$$

3. Confidence intervals for estimated sample sizes.

A single determination of the sample size using the estimates from the LMM alone does not provide any quantification of uncertainty in these measures. Relying solely on this point estimate for a clinical trial may risk underpowering (or overpowering) the study. For this reason, it is informative to construct confidence intervals around estimated sample sizes to provide a guide to the precision of the estimates. Since the sampling distribution of sample size estimates is complex and not readily amenable to approximation with explicit algebraic formulae, the bootstrap[7] can be used to do this. The other advantage of utilising bootstrap is that it provides additional robustness for the confidence interval if the assumptions of the linear mixed models used in the analysis do not hold exactly.

Here we constructed non-parametric bias-corrected and accelerated (BCa) confidence intervals from 5000 bootstrap samples for each of our sample size estimates. The confidence intervals were constructed on the “effect sizes”, i.e. the ratio of the target treatment effect to its standard error for a two-person trial. For example, for the ANCOVA sample size calculation above the effect size would be $d/\left( 2(\sigma_{F}^{2}-\sigma_{BF}^{2}/\sigma_{B}^{2}) \right)^{1/2}$. The distribution of estimated effect sizes is likely to be more symmetric than that of estimated sample sizes and so confidence intervals calculated on this scale are likely to have better coverage properties. The confidence intervals were then transformed to the sample size scale.

4. Computational details

For computational convenience, we used an adapted version of the Stata package slopepower[8]. Slopepower incorporates the two stages of first fitting the LMM to observational data and then using the model estimates to calculate a sample size for a postulated trial. While slopepower uses the more general matrix algebra approach found in[1,2], for the simple case of a baseline measure and a single follow-up measure it is equivalent to using ANCOVA to calculate the sample size. The adaptations to slopepower include incorporating adjustment for TIV, allowing for analysis of outcomes based on direct measures of change, and bootstrapping the effect size.

The analysis of the observational data requires linear mixed models to be fitted to non-carriers and carriers separately, although for convenience this was done using a single joint model with all parameters (including random effects and residuals) allowed to differ between carriers and non-carriers.

The LMM was considered to have failed to converge if it either produced an error during the fitting process or resulted in an estimate of the correlation between slope and intercept with an absolute value of greater than 0.99, as this suggests that the parameter estimate is on the boundary of the parameter space.

Some checks were carried out before including the specific outcomes in the sample size analysis. First, estimates for the difference in slope between carriers and non-carriers were obtained from the LMMs specified in (1) and (2) for those variables reflecting neurodegeneration. When the ratio between the estimate of the slope difference and the standard error of this estimate (effectively a z-score) was less than 2.5, then this outcome was excluded from sample size analysis. This is because the difference in slopes between carriers and non-carriers is not highly significant, and so it would be unlikely to be a suitable candidate for intervention in a future trial where the target therapeutic effect is based on a 25% change in slope. In addition, there would likely be a large enough proportion of bootstrap samples that had effect sizes with a flipped sign (*i.e.* less disease in carriers than non-carriers), such that the confidence intervals for the effect size would span zero. This would mean that the upper limit of the 95% CI for the sample size could not be estimated. The check on the ratio between the estimate of the slope difference and its standard error was applied to both the model fitted to the entire trial eligible sample with baseline global CDR=0-1, as well as to the subset of participants with a global CDR score of 0 at baseline.

REFERENCES

[1]	Frost C, Mulick A, Scahill RI, Owen G, Aylward E, Leavitt BR, et al. Design optimization for clinical trials in early-stage manifest Huntington’s disease. Movement Disorders 2017;32:1610–9. https://doi.org/10.1002/mds.27122.

[2]	Frost C, Kenward MG, Fox NC. Optimizing the design of clinical trials where the outcome is a rate . Can estimating a baseline rate in a run-in period increase efficiency? Stat Med 2008;27:3717–31. https://doi.org/10.1002/sim.

[3]	Frost C, Kenward MG, Fox NC. The analysis of repeated “direct” measures of change illustrated with an application in longitudinal imaging. Stat Med 2004;23:3275–86. https://doi.org/10.1002/sim.1909.

[4]	Cash DM, Frost C, Iheme LO, Ünay D, Kandemir M, Fripp J, et al. Assessing atrophy measurement techniques in dementia: Results from the MIRIAD atrophy challenge. Neuroimage 2015;123:149–64. https://doi.org/10.1016/j.neuroimage.2015.07.087.

[5]	Dawson JD. Sample Size Calculations Based on Slopes and Other Summary Statistics. Biometrics 1998;54:323–30.

[6]	Frison L, Pocock SJ. Repeated measures in clinical trials: analysis using mean summary statistics and its implications for design. Stat Med 1992;11:1685–704.

[7]	Efron B, Tibshirani RJ. An introduction to the bootstrap. vol. 57. CRC press; 1994.

[8]	Nash S, Morgan KE, Frost C, Mulick A. Power and sample-size calculations for trials that compare slopes over time: Introducing the slopepower command: Stata J 2021;21:575–601. https://doi.org/10.1177/1536867X211045512.

**Supplemental Table 1** Estimated outcome measures at baseline and at four-year follow-up (with 95% confidence intervals) for carriers and non-carriers, based on the linear mixed effects model of participants in the observational studies. These estimates form the basis of the subsequent sample size estimates. For models where the outcome was transformed, the estimates have been back-transformed to the original scale. Outcomes with blank cells are where the linear mixed model failed to converge or there was insufficient evidence of change in slope between carriers and non-carriers.

|  | | **Non-Carrier** | | **Carrier (CDR=0)** | | **Carrier (CDR=0-1)** | |
| --- | --- | --- | --- | --- | --- | --- | --- |
| Modality | Outcome | Baseline | Year 4 | Baseline | Year 4 | Baseline | Year 4 |
| **COG** | MMSE | 29.2 (29.0, 29.4) | 29.3 (29.1, 29.5) | 28.9 (28.6, 29.2) | 27.9 (27.2, 28.6) | 27.1 (26.5, 27.7) | 23.0 (21.6, 24.4) |
|  | CDR SOB | 0.04 (-0.01, 0.09) | 0.08 (0.03, 0.13) |  |  | 1.03 (0.72, 1.35) | 4.21 (3.20, 5.23) |
|  | Cognitive Composite 1 | -0.03 (-0.14, 0.08) | 0.12 (0.00, 0.23) | -0.23 (-0.37, -0.098) | -0.39 (-0.59, -0.19) | -0.774 (-0.94, -0.61) | -1.47 (-1.78, -1.17) |
| **MRI (ml)** | Brain Volume | 1130 (1120, 1130) | 1120 (1110, 1130) | 1120 (1110, 1130) | 1110 (1090, 1120) | 1110 (1100, 1120) | 1080 (1070, 1090) |
|  | Ventricular Volume | 17.7 (16.4, 19.1) | 18.5 (17.1, 20.0) | 17.9 (16.1, 19.8) | 20.7 (18.2, 23.5) | 21.0 (19.2, 22.9) | 27.7 (24.5, 31.3) |
|  | Hippocampal Volume | 6.43 (6.31, 6.55) | 6.39 (6.27, 6.51) | 6.38 (6.27, 6.50) | 6.22 (6.08, 6.37) | 6.11 (5.98, 6.24) | 5.79 (5.63, 5.95) |
|  | Post. Cing. Volume | 9.98 (9.78, 10.2) | 9.89 (9.70, 10.1) | 9.66 (9.43, 9.90) | 9.39 (9.11, 9.67) | 9.45 (9.28, 9.63) | 8.94 (8.71, 9.18) |
|  | Precuneus Volume | 20.1 (19.7, 20.6) | 19.9 (19.5, 20.3) | 19.7 (19.3, 20.1) | 18.9 (18.4, 19.5) | 18.8 (18.4, 19.3) | 17.4 (16.8, 18.1) |
| **PIB** | Inf. Par. SUVR | 0.91 (0.87, 0.95) | 0.90 (0.86, 0.95) | 1.46 (1.29, 1.66) | 1.67 (1.45, 1.94) | 1.64 (1.49, 1.81) | 1.90 ( 1.70, 2.12) |
|  | Inf. Temp. SUVR | 0.89 (0.86, 0.92) | 0.88 (0.85, 0.91) | 1.29 (1.15, 1.44) | 1.50 (1.31, 1.72) | 1.41 (1.29, 1.54) | 1.67 ( 1.50, 1.87) |
|  | Mid. Temp.SUVR | 0.82 (0.79, 0.85) | 0.80 (0.77, 0.83) | 1.22 (1.08, 1.38) | 1.42 (1.24, 1.63) | 1.34 (1.21, 1.48) | 1.57 (1.41, 1.75) |
|  | Post. Cing. SUVR | 1.21 (1.17, 1.24) | 1.23 (1.19, 1.27) | 1.86 (1.63, 2.11) | 2.09 (1.81, 2.41) | 2.16 (1.95, 2.39) | 2.39 (2.13, 2.67) |
|  | Precuneus SUVR | 1.13 (1.09, 1.17) | 1.12 (1.08, 1.17) | 1.98 (1.72, 2.28) | 2.29 (1.93, 2.71) | 2.38 (2.13, 2.66) | 2.72 (2.39, 3.09) |
|  | Cortical SUVR | 1.04 (1.01, 1.06) | 1.02 (0.99, 1.05) | 1.68 (1.48, 1.91) | 1.90 (1.64, 2.21) | 1.95 (1.76, 2.16) | 2.18 (1.94, 2.45) |
| **FDG** | Banks STS SUVR | 1.22 (1.19, 1.24) | 1.20 (1.17, 1.23) |  |  | 1.16 (1.13, 1.18) | 1.05 (1.01, 1.09) |
|  | Inf. Par. SUVR | 1.18 (1.16, 1.21) | 1.17 (1.14, 1.20) | 1.17 (1.14, 1.20) | 1.11 (1.07, 1.16) | 1.08 (1.05, 1.12) | 0.98 (0.93, 1.03) |
|  | Post. Cing. SUVR | 1.26 (1.23, 1.28) | 1.24 (1.22, 1.27) | 1.23 (1.20, 1.26) | 1.22 (1.18, 1.26) | 1.18 (1.16, 1.21) | 1.13 (1.10, 1.17) |
|  | Precuneus SUVR | 1.37 (1.35, 1.40) | 1.36 (1.33, 1.39) |  |  | 1.24 (1.21, 1.28) | 1.12 (1.07, 1.19) |
|  | Hippocampus SUVR | 0.89 (0.88, 0.91) | 0.89 (0.87, 0.90) | 0.89 (0.87, 0.90) | 0.89 (0.87, 0.91) | 0.87 (0.85, 0.88) | 0.85 (0.83, 0.87) |
|  | Cortical SUVR | 1.20 (1.18, 1.23) | 1.18 (1.16, 1.21) |  |  | 1.15 (1.12, 1.17) | 1.08 (1.05, 1.12) |
| **CSF (pg/ml)** | XMAP tTau | 52.7 (47.5, 58.5) | 52.0 (46.7, 58.0) | 88.0 (75.9, 102) | 85.4 (73.5, 99.1) | 113 (100, 127) | 109 (97.5, 123) |
|  | XMAP tTau Long | 43.5 (39.2, 48.3) | 43.3 (38.7, 48.5) | 66.8 (56.5, 79.0) | 71.7 (60.0, 85.7) | 85.1 (75.1, 96.4) | 86.6 (75.9, 98.7) |
|  | XMAP Aβ42 Long | 518 ( 470, 570) | 499 ( 449, 554) | 313 ( 268, 367) | 257 ( 215, 306) | 257 ( 230, 288) | 204 (179, 233) |
|  | XMAP pTau181 Long | 21.3 (19.7, 23.0) | 15.5 (13.9, 17.3) |  |  | 42.1 (36.4, 48.6) | 33.5 (29.4, 38.2) |
|  | Lumipulse Aβ40 | 8770 (8150, 9450) | 8370 (7760, 9030) | 8290 (7630, 9000) | 7190 (6570, 7860) | 8320 (7850, 8830) | 7090 (6620, 7580) |
|  | Lumipulse Aβ42 | 787 (728, 850) | 755 ( 697, 818) | 512 ( 459, 570) | 411 ( 366, 461) | 435 ( 401, 472) | 351 ( 321, 384) |
|  | Lumipulse tTau | 249 ( 230, 270) | 244 ( 224, 266) | 410 ( 356, 472) | 444 ( 384, 514) | 523 ( 468, 584) | 575 ( 512, 647) |
|  | Lumipulse pTau181 | 26.6 (24.2, 29.1) | 25.3 (23.1, 27.8) | 56.2 (46.9, 67.3) | 61.2 (50.7, 73.9) | 78.7 (68.4, 90.4) | 85.8 (74.2, 99.3) |
|  | Lumipulse Aβ42/40 | 0.090 (0.087, 0.093) | 0.090 (0.087, 0.094) | 0.062 (0.057, 0.067) | 0.057 (0.053, 0.062) | 0.052 (0.049, 0.056) | 0.050 (0.047, 0.053) |
|  | Lumipulse ptau181/Aβ42 | 0.034 (0.032, 0.036) | 0.034 (0.031, 0.036) | 0.109 (0.087, 0.138) | 0.151 (0.118, 0.193) | 0.181 (0.150, 0.218) | 0.242 (0.198, 0.295) |

**Supplemental Table 2** Sample size estimates (with 95% bootstrap confidence intervals, bias corrected and accelerated) per arm needed to detect a 50% reduction in the annual rate of change in a four-year trial, assuming 40% dropout after four years. Outcomes with blank cells are where the linear mixed model failed to converge. Results in italics indicate that the number of bootstrap samples where the model failed to converge is greater than 1%, so these confidence intervals should be treated with caution.

| Modality | Outcome | CDR=0 | CDR=0-1 |
| --- | --- | --- | --- |
| COG | CDR SOB |  | *161 (102,291)* |
|  | MMSE | *878 (353,>10000)* | 204 (123,411) |
|  | Cog Composite | *326 (157,1074)* | 104 (68,192) |
| MRI | Brain Volume |  | *118 (59,338)* |
|  | Brain BSI | 727 (318,2398) | 229 (148,325) |
|  | Ventricular Volume | 246 (120,980) | 77 (50,137) |
|  | Ventricle BSI | 410 (218,941) | 150 (102,198) |
|  | Hippocampal Volume | 338 (131,2096) | 142 (88,279) |
|  | Post. Cing. Volume |  | 217 (120,626) |
|  | Precuneus Volume | 649 (254,>10000) | 137 (80,284) |
| FDG | Cortical SUVR |  | *256 (100,1208)* |
|  | Inf. Par. SUVR |  | *146 (71,428)* |
|  | Precuneus SUVR |  | *121 (56,368)* |
|  | Banks STS SUVR |  | *115 (56,268)* |

Supplemental Figure 1 Estimated measures from all outcomes at baseline and at four-year follow-up (means and 95% confidence intervals) for carriers and non-carriers. These estimates are based on the fitted parameters from fitting the linear mixed effects model to participants in the observational studies that match the trial eligibility criteria. These estimates form the basis of the subsequent sample size estimates. All estimates have been back-transformed, when necessary, in order to plot the estimated outcome measures in the original scale.

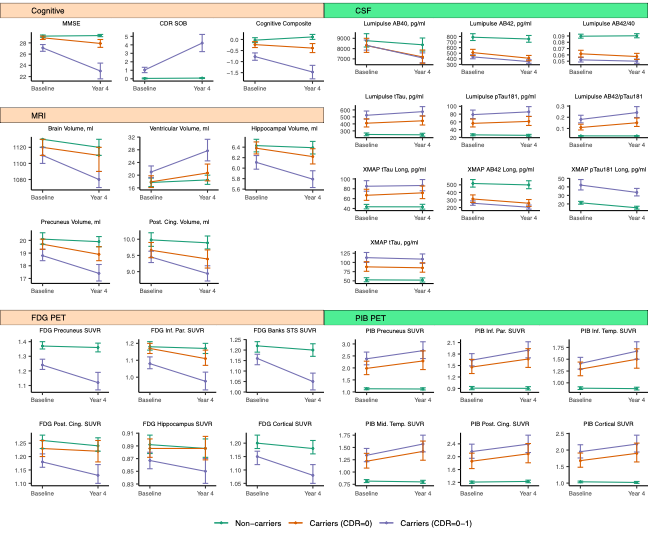
